# ‘Just part of the job’ – understanding work-related injuries and safety culture in companion animal veterinary practices

**DOI:** 10.1101/2025.05.28.25328510

**Authors:** John S.P. Tulloch, Imogen Schofield, Rebecca Jackson, Martin Whiting

## Abstract

**Objectives:** To examine the prevalence and types of work-related injuries in companion animal practices, explore the context of their occurrence, and the behaviours of injured persons.

**Methods:** A mixed-methods analysis of a cross-sectional online survey of UK employees of a consolidated group of veterinary practices.

**Results:** Of 647 respondents, 77.6% experienced a work-related injury during their career. In the previous year, 60.2% of veterinary nurses and 58.3% of veterinarians were injured, most frequently in consultation rooms, prep areas, kennels, and reception. Animal-related injuries were the most prevalent injury type. Injuries frequently occurred during cat restraint, anaesthetic recovery, and clinical examinations. Needlestick injuries made up 15.8% of veterinary injuries. 16.3% of injured nurses and 19.4% of injured vets attended hospital. 34.3% of nurses, and 25.1% of vets, needed more than a week to recover from their injuries. Fewer than 10% took time off work, often due to a sense of duty, the ability to manage a reduced workload, or simply wanting to “get on with it.” Most injuries to vets went unreported, due to perceived time pressures or the belief that the injury was minor. Around half adjusted their behaviour post-injury, becoming more cautious or changing handling techniques.

**Clinical significance:** This study reveals a high rate of work-related injuries in companion animal practices. A culture of presenteeism and blame often downplays these risks, hindering safety. To reduce injuries, a shift towards shared responsibility and reflective learning is needed, driven by strong leadership and open communication.

## Introduction

The veterinary industry has some of the highest levels of work-related injuries (US Bureau of Labor Statistics 2023). In the United Kingdom (UK) around two-thirds of veterinarians are injured annually (Mills 2019). It has been estimated that 95% of companion animal veterinarians have been injured within the last five years (Epp & Waldner 2012), with between 26-34% being injured in the last 12 months (Fritschi *et al*. 2006; Johnson & Fritschi 2024).

Companion animal veterinary practices present various workplace injury risks, primarily due to animals, handling of sharp instruments, and exposure to hazardous substances (Epp & Waldner 2012). Animal-related injuries (cat bites and scratches, and dog bites) are the most prevalent cause of injury (Johnson & Fritschi 2024; Tulloch *et al*. 2023; Voss *et al*. 2023), often occurring during the restraint or clinical examination or administration of drugs to said animal (Tulloch *et al*. 2023). Many of these animal-related injuries will require medical attention, with varied estimates of the number requiring hospital treatment (4-16%) (Lucas *et al*. 2009; Tulloch *et al*. 2023). Musculoskeletal injuries can occur from lifting heavy animals or maintaining awkward postures during procedures, with 46-49% of veterinary surgeons receiving one annually (Epp & Waldner 2012; Fritschi *et al*. 2006). Needlestick injuries and exposure to anaesthetic gases, disinfectants, or medications pose additional health risks (Epp & Waldner 2012; Johnson & Johnstone 2025). It has been estimated that 57-65% of veterinarians receive a needlestick annually (Epp & Waldner 2012; Johnson & Fritschi 2024).

Despite there being literature describing injuries to veterinarians there is little describing the context of those injuries, their consequences, nor the health-seeking or reporting behaviours of the injured person (IP). There is also scant information about other roles being injured in a companion animal veterinary practice (i.e. veterinary nurses, receptionists, animal care assistants). A strong safety culture within a workplace, defined by shared values, attitudes, and behaviours regarding workplace safety, plays a crucial role in mitigating risks and preventing injuries. It appears that there is some shared culture between veterinary roles as they have a consistent definition of a work-related injury being one causing ‘physical trauma with pain’, however one in ten also incorrectly believe that preceding events must be an accident for something to be defined as an injury (Furtado *et al*. 2024). In UK companion animal practices injury prevalence, health seeking, and reporting behaviours remain underexplored. Understanding the relationship between safety culture and work-related injuries is essential for developing targeted interventions, improving training programs, and fostering a safer working environment for veterinary professionals.

This paper aims to examine the prevalence and types of work-related injuries in companion animal practices, explore the context of their occurrence, and the behaviours of injured persons. The study seeks to provide actionable insights for enhancing occupational health in veterinary settings.

## Materials and Methods

An online cross-sectional survey was designed and distributed to all UK employees of CVS Group plc, working in companion animal clinical practices. The survey was distributed through weekly emails and was open for three months between 6^th^ December 2022 and 6^th^ March 2023. The survey was piloted with a small group of CVS employees. To enhance recruitment, an incentive of a year’s supply of tea, coffee, and biscuits, for the practices of five randomly selected respondents was provided.

Respondents were asked about personal demographics (i.e. sex, age, nationality) and details of their job role, number of hours worked, and whether they perform clinical work. Then about whether they had acquired a work-related injury during their career in the veterinary sector. If they had, they were presented with a series of questions about their most recent injury and their most severe injury. These included questions about injury context, medical consequences, resultant behaviour change, time off work, and reporting to their employer.

Demographic characteristics of respondents were described. Overall injury prevalence was calculated and stratified by job role. Logistic regression was performed to identify any association between demographic variables and ever receiving an injury. Variables taken forward for multivariable analysis were selected through substantive knowledge and statistical significance (i.e. where p<=0.3). Hosmer-Lemeshow tests were performed to assess goodness of fit, and analysis of a correlation matrix of independent variables was used to assess collinearity.

The data was subsequently stratified from the remainder of the analysis into the following groups; administrative, receptionists, animal care assistants (ACA), veterinary nurses (VN), and veterinary surgeons (VS). The types of injury acquired were described and it was recorded whether they were reportable to the Health and Safety Executive (HSE). In the UK, under the Reporting of Injuries, Diseases and Dangerous Occurrences Regulation 2013 (RIDDOR), an employer legally must report work-related accidents that result in specific injuries to the HSE (Health & Safety Executive 2013). These injuries include any that leave them incapacitated for more than seven days, fractures other than to digits, loss of consciousness, among others. We performed more focused analysis on animal-related injuries, and the procedures occurring at the point of injury, as we anticipated that these would represent the majority of injuries to clinical staff.

The analysis of questions relating to the context of injury, attitude of injured person, and reporting culture were carried out descriptively. Open text questions were analysed using an iterative thematic analysis (Vears & Gillam 2022). Initially, the responses were read through with the researchers making note of any initial impressions; secondly, the researchers carried out initial “coding”, by reading items individually and labelling important concepts. As more text was incorporated into the analysis, codes were refined, combined, or renamed to more accurately represent the information conveyed by respondents. Eventually, codes could be grouped into overall categories or “themes”.

Data masking was used to protect personally identifiable information of respondents, this was performed through aggregation of data or redacting of quotes. The study received ethical approval from the University of Liverpool Veterinary Research Ethics Committee (VREC1256).

## Results

Six hundred and forty-seven individuals consented to take part in the survey. This represents a response rate of 10.5%. VS respondents were representative of the UK profession in terms of age (median national age 30-39) and ethnicity (3.5% ethnic minority groups nationally) (Table 1) (Robinson *et al*. 2019a). However, female respondents were over-represented (59% of veterinarians are female nationally). VNs were broadly representative in terms of sex (96.8% female nationally), age (median age of 30-39 nationally), and ethnicity (1.9% ethnic minority groups nationally) (Robinson *et al*. 2019b). National demographics of companion animal ACAs, administrators, and receptionist are currently not available.

**Table 1:**
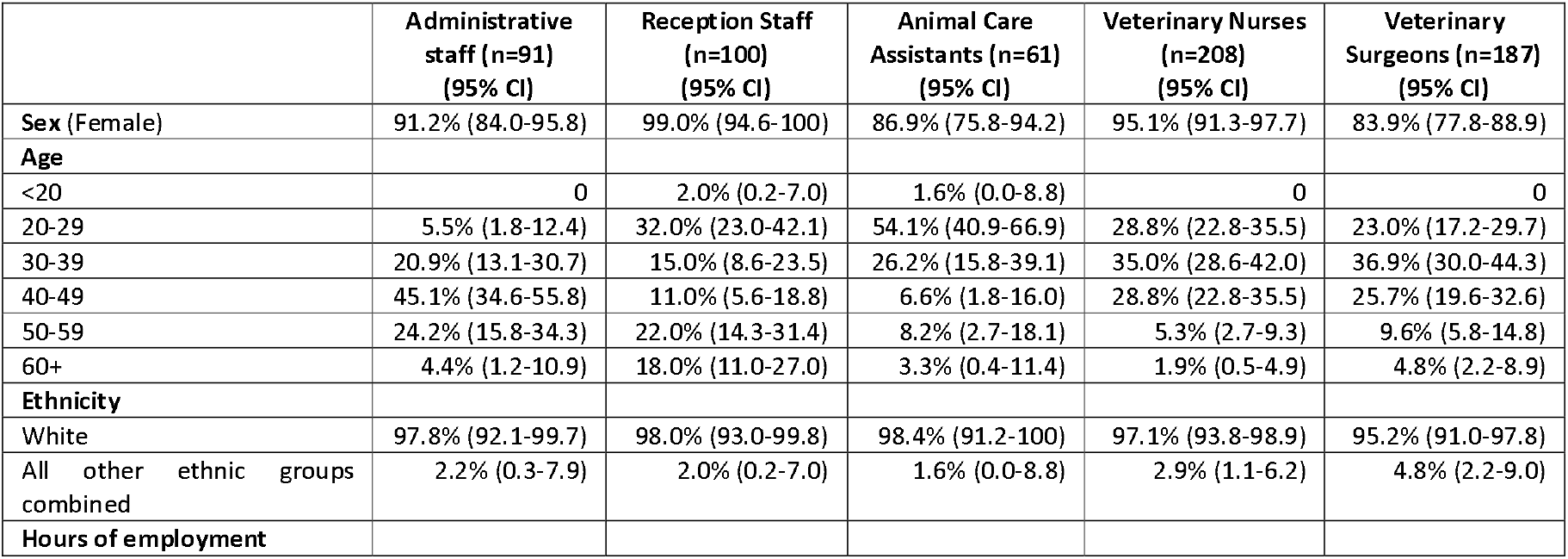
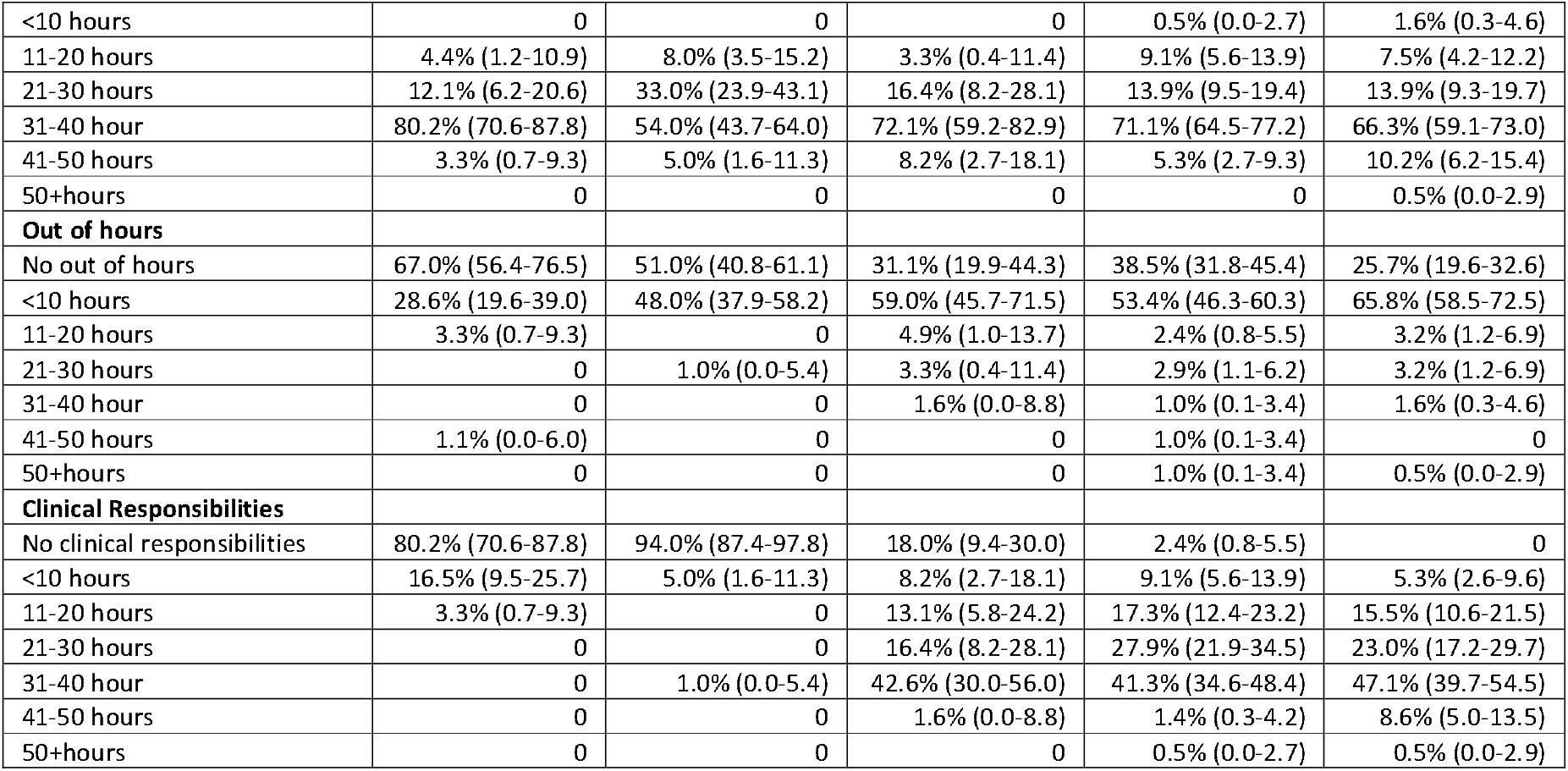
Demographics of respondents to a survey on veterinary work-related injuries in companion animal practices.

|  | Administrative staff (n=91)<br>(95% CI) | Reception Staff (n=100)<br>(95% CI) | Animal Care Assistants (n=61)<br>(95% CI) | Veterinary Nurses (n=208)<br>(95% CI) | Veterinary Surgeons (n=187)<br>(95% CI) |
| --- | --- | --- | --- | --- | --- |
| **Sex (Female)** | 91.2% (84.0-95.8) | 99.0% (94.6-100) | 86.9% (75.8-94.2) | 95.1% (91.3-97.7) | 83.9% (77.8-88.9) |
| **Age** |  |  |  |  |  |
| <20 | 0 | 2.0% (0.2-7.0) | 1.6% (0.0-8.8) | 0 | 0 |
| 20-29 | 5.5% (1.8-12.4) | 32.0% (23.0-42.1) | 54.1% (40.9-66.9) | 28.8% (22.8-35.5) | 23.0% (17.2-29.7) |
| 30-39 | 20.9% (13.1-30.7) | 15.0% (8.6-23.5) | 26.2% (15.8-39.1) | 35.0% (28.6-42.0) | 36.9% (30.0-44.3) |
| 40-49 | 45.1% (34.6-55.8) | 11.0% (5.6-18.8) | 6.6% (1.8-16.0) | 28.8% (22.8-35.5) | 25.7% (19.6-32.6) |
| 50-59 | 24.2% (15.8-34.3) | 22.0% (14.3-31.4) | 8.2% (2.7-18.1) | 5.3% (2.7-9.3) | 9.6% (5.8-14.8) |
| 60+ | 4.4% (1.2-10.9) | 18.0% (11.0-27.0) | 3.3% (0.4-11.4) | 1.9% (0.5-4.9) | 4.8% (2.2-8.9) |
| **Ethnicity** |  |  |  |  |  |
| White | 97.8% (92.1-99.7) | 98.0% (93.0-99.8) | 98.4% (91.2-100) | 97.1% (93.8-98.9) | 95.2% (91.0-97.8) |
| All other ethnic groups combined | 2.2% (0.3-7.9) | 2.0% (0.2-7.0) | 1.6% (0.0-8.8) | 2.9% (1.1-6.2) | 4.8% (2.2-9.0) |
| **Hours of employment** |  |  |  |  |  |
| <10 hours | 0 | 0 | 0 | 0.5% (0.0-2.7) | 1.6% (0.3-4.6) |
| 11-20 hours | 4.4% (1.2-10.9) | 8.0% (3.5-15.2) | 3.3% (0.4-11.4) | 9.1% (5.6-13.9) | 7.5% (4.2-12.2) |
| 21-30 hours | 12.1% (6.2-20.6) | 33.0% (23.9-43.1) | 16.4% (8.2-28.1) | 13.9% (9.5-19.4) | 13.9% (9.3-19.7) |
| 31-40 hours | 80.2% (70.6-87.8) | 54.0% (43.7-64.0) | 72.1% (59.2-82.9) | 71.1% (64.5-77.2) | 66.3% (59.1-73.0) |
| 41-50 hours | 3.3% (0.7-9.3) | 5.0% (1.6-11.3) | 8.2% (2.7-18.1) | 5.3% (2.7-9.3) | 10.2% (6.2-15.4) |
| 50+hours | 0 | 0 | 0 | 0 | 0.5% (0.0-2.9) |
| **Out of hours** |  |  |  |  |  |
| No out of hours | 67.0% (56.4-76.5) | 51.0% (40.8-61.1) | 31.1% (19.9-44.3) | 38.5% (31.8-45.4) | 25.7% (19.6-32.6) |
| <10 hours | 28.6% (19.6-39.0) | 48.0% (37.9-58.2) | 59.0% (45.7-71.5) | 53.4% (46.3-60.3) | 65.8% (58.5-72.5) |
| 11-20 hours | 3.3% (0.7-9.3) | 0 | 4.9% (1.0-13.7) | 2.4% (0.8-5.5) | 3.2% (1.2-6.9) |
| 21-30 hours | 0 | 1.0% (0.0-5.4) | 3.3% (0.4-11.4) | 2.9% (1.1-6.2) | 3.2% (1.2-6.9) |
| 31-40 hours | 0 | 0 | 1.6% (0.0-8.8) | 1.0% (0.1-3.4) | 1.6% (0.3-4.6) |
| 41-50 hours | 1.1% (0.0-6.0) | 0 | 0 | 1.0% (0.1-3.4) | 0 |
| 50+hours | 0 | 0 | 0 | 1.0% (0.1-3.4) | 0.5% (0.0-2.9) |
| **Clinical Responsibilities** |  |  |  |  |  |
| No clinical responsibilities | 80.2% (70.6-87.8) | 94.0% (87.4-97.8) | 18.0% (9.4-30.0) | 2.4% (0.8-5.5) | 0 |
| <10 hours | 16.5% (9.5-25.7) | 5.0% (1.6-11.3) | 8.2% (2.7-18.1) | 9.1% (5.6-13.9) | 5.3% (2.6-9.6) |
| 11-20 hours | 3.3% (0.7-9.3) | 0 | 13.1% (5.8-24.2) | 17.3% (12.4-23.2) | 15.5% (10.6-21.5) |
| 21-30 hours | 0 | 0 | 16.4% (8.2-28.1) | 27.9% (21.9-34.5) | 23.0% (17.2-29.7) |
| 31-40 hours | 0 | 1.0% (0.0-5.4) | 42.6% (30.0-56.0) | 41.3% (34.6-48.4) | 47.1% (39.7-54.5) |
| 41-50 hours | 0 | 0 | 1.6% (0.0-8.8) | 1.4% (0.3-4.2) | 8.6% (5.0-13.5) |
| 50+hours | 0 | 0 | 0 | 0.5% (0.0-2.7) | 0.5% (0.0-2.9) |

More than three quarters of all staff had acquired a work-related injury in the veterinary industry during their career (77.6%). This was highest for VS (93.6%), followed by VNs (92.8%), ACAs (75.4%), administrators (56.0%) and receptionists (37.0%). Veterinarians and VNs reported the highest frequency of injuries (Fig 1).

**Figure 1:**
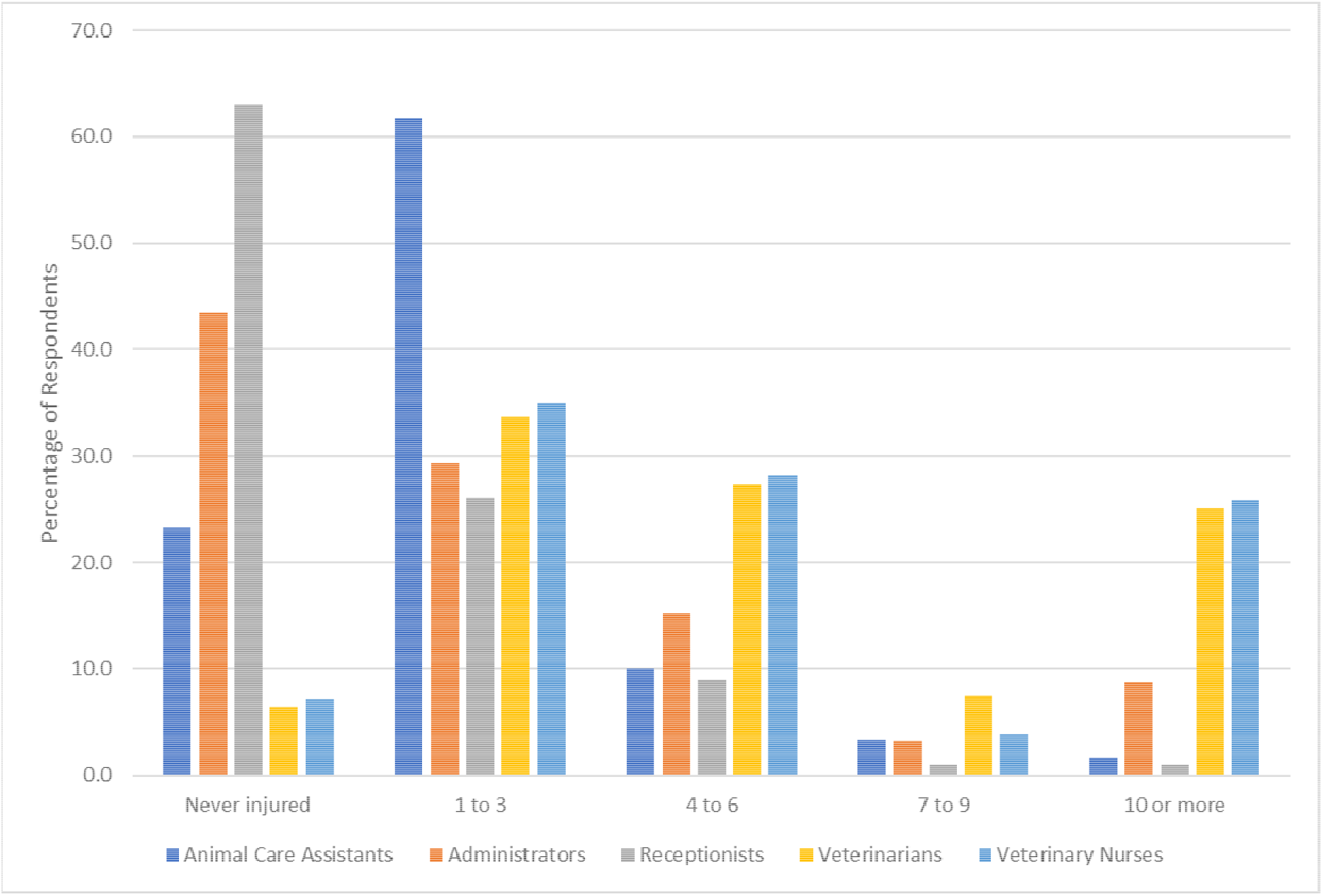
Frequency of work-related injuries over a career in companion animal practices.

Univariable analysis to identify which demographic variables were associated with a respondent having ever acquired a veterinary work-related injury identified the following significant variables; sex, role, hours worked, out of hours worked, and clinical hours worked (Table 2). When taking forward variables for multivariable analysis, “clinical hours” was removed as there was strong collinearity with “role”. The multivariable analysis showed that role and hours worked were the variables with significance. All roles had higher odds of having acquired an injury, when adjusted for sex, hours worked, and OOH worked, than receptionists. The roles with the highest odds of injury were VSs and VNs. Only those that worked between 21-40 hours a week had higher odds of injury compared to those working less than 21 hours, those working more than 40 hours had similar injury odds to those working the fewest hours.

**Table 2:** Univariable and multivariable logistic regression exploring demographic factors associated with acquiring a veterinary work-related injury in companion animal practices.

|  | Univariable analysis |  | Multivariable analysis |  |
| --- | --- | --- | --- | --- |
|  | Odds Ratio<br>(95% CI) | p-value | Odds Ratio<br>(95% CI) | p-value |
| **Sex (Ref: Female)** | 2.2 (1.0-5.4) | 0.06 | 1.2 (0.5-3.3) | 0.63 |
| **Age (Ref: <=29 years old)** |  |  |  |  |
| 30-39 | 1.4 (0.9-2.3) | 0.19 |  |  |
| 40-49 | 1.3 (0.8-2.2) | 0.34 |  |  |
| 50-59 | 0.8 (0.5-1.5) | 0.53 |  |  |
| 60+ | 0.8 (0.4-1.7) | 0.50 |  |  |
| **Ethnicity (Ref: White)** |  |  |  |  |
| All other ethnic groups combined | 2.7 (0.8-17.0) | 0.19 |  |  |
| **Role (Ref: Receptionist)** |  |  |  |  |
| Animal Care Assistant | 5.2 (2.6-10.9) | <0.001 | 4.9 (2.4-10.5) | <0.001 |
| Administrator | 2.2 (1.2-3.9) | <0.01 | 2.0 (1.1-3.7) | 0.03 |
| Veterinary Nurse | 21.9 (11.6-42.9) | <0.001 | 24.1 (12.2-50.4) | <0.001 |
| Veterinary Surgeon | 24.8 (12.6-52.7) | <0.001 | 28.5 (13.6-64.2) | <0.001 |
| **Weekly hours worked**<br>**(Ref: <=20hours)** |  |  |  |  |
| 21-30 | 1.4 (0.7-2.9) | 0.32 | 4.1 (1.6-10.4) | <0.01 |
| 31-40 | 2.3 (1.2-4.2) | 0.01 | 4.4 (1.9-9.9) | <0.001 |
| 41+ | 1.6 (0.7-4.1) | 0.28 | 1.9 (0.6-5.8) | 0.28 |
| **Weekly ‘out of hours’ worked**<br>**(Ref: 0 hours)** |  |  |  |  |
| >0-10 | 1.7 (1.2-2.5) | <0.01 | 0.96 (0.6-1.6) | 0.87 |
| 11-20 | 3.1 (0.8-19.7) | 0.14 | 1.9 (0.4-13.2) | 0.45 |
| >=21 | 5.1 (1.5-32.2) | 0.03 | 1.9 (0.5-13.0) | 0.44 |
| **Weekly clinical hours worked**<br>**(Ref: 0 hours)** |  |  |  |  |
| >0-10 | 10.3 (4.5-27.9) | <0.001 |  |  |
| 11-20 | 6.9 (3.6-14.2) | <0.001 |  |  |
| 21-30 | 27.3 (11.7-80.0) | <0.001 |  |  |
| 31-40 | 16.0 (9.0-30.2) | <0.001 |  |  |
| 41+ | 5.8 (2.1-20.7) | <0.01 |  |  |
| **Knowledge of Health and Safety Protocol**<br>**(Ref: No knowledge)** | 1.38 (0.8-2.2) | 0.19 |  |  |

The last 12 months annual work-related injury prevalence for administrators was 12.1% (95%CI 6.2-20.6), for receptionists 21.0% (13.5-30.3), for ACAs 62.3% (49.0-74.4), for VNs 60.6% (53.4-67.3), and for VS 58.3% (50.9-65.4). Over 90% of all staff were injured within the practice. The most prevalent areas for injuries were the consult room, the prep room, the ward/kennels area, and reception (Table 3, Table S1).

**Table 3:**
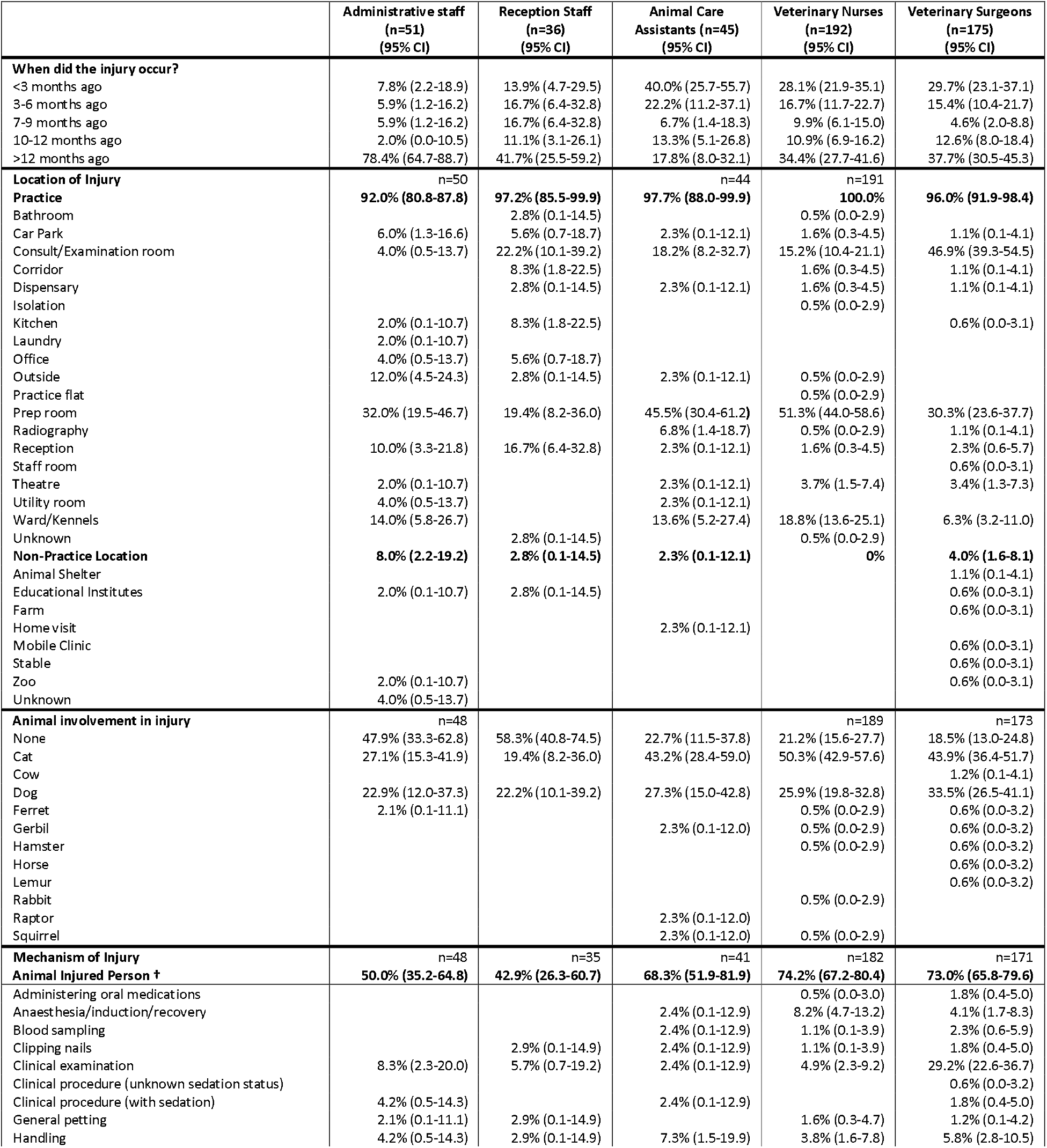
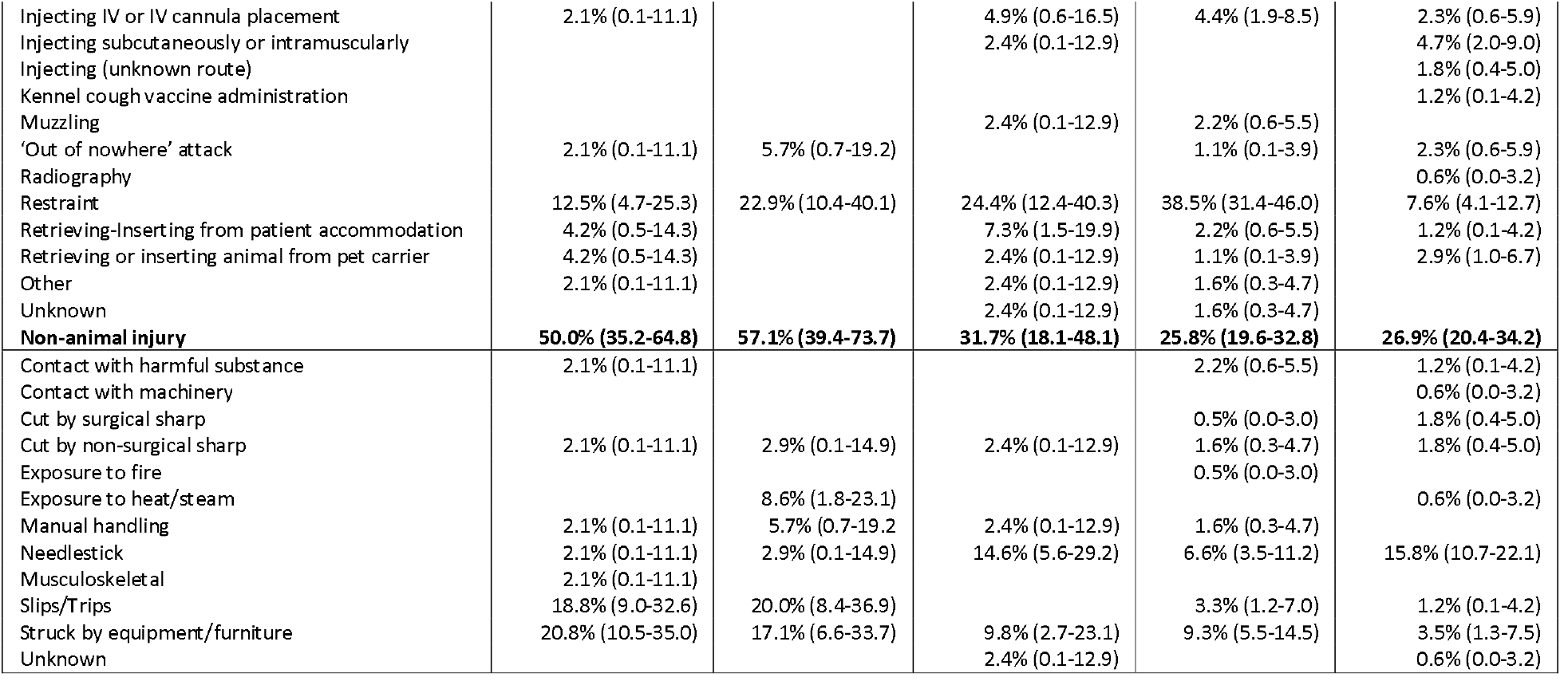
Context of recent veterinary work-related injuries in companion animal practices. †Will be lower than animal involvement in injury. For example, sharps are not classified as having animal involvement, but if occurred during surgery an individual may say an animal was involved.

### Mechanism of Injury

Administrative staff were injured in highly varied ways, with no clear discernible patterns. The top mechanisms of injury were ‘struck by equipment or furniture’, ‘slips/trips’ and ‘restraint of an animal’. Receptionists’ injuries primarily did not involve animals; however, the top injury mechanism was ‘restraint of an animal’ followed by ‘slips/trips’ and ‘struck by equipment/furniture.’ When animals were involved with injuries to these job roles, they were almost exclusively cats and dogs. The majority of ACAs injuries involved animals; cats were the most prevalent animal involved followed by dogs. Around a quarter of all injuries involved the restraint of an animal, these all involved the restraint for a veterinarian to carry out a procedure. There was no obvious trend for procedure type, however cats were involved in 80% of these incidences. The only other injury mechanism with a prevalence higher than 10% were needlestick injuries.

The majority of injuries to VNs were animal-related with around half involving cats, with dogs being the next most prevalent animal involved. Injuries mainly occurred when the VN was restraining an animal, in 83.0% of recent and 82.1% of severe injuries these were cats with the remainder being dogs. The top three reasons for cats being restrained were for blood samples, injections, and clinical examinations. The next most prevalent injury mechanism was during an animal’s ‘anaesthesia/induction/recovery’, in 73.3% of recent and 76.5% of severe injuries this led to a bite, and in 46.7% and 47.0% of cases it was stated that the ‘animal suddenly woke up’. Similarly, the majority of injuries to veterinarians were by animals, with cats being more prevalent in the most recent injuries, and dogs being more prevalent in the most severe injuries. Clinical examinations were the most common activity occurring at the point of injury; 51.8% recent & 40.9% severe were examinations to the head, anal examinations (11.1% recent, 18.2% severe), chest examinations (11.1% recent, 9.1% severe) and abdominal examinations (11.1% recent, 4.5% severe). Needlesticks were the next most prevalent injury mechanism, representing almost 1 in 6 of all recent injuries to veterinarians.

Needlesticks represented a large burden of injury across all roles (Table 4). Many needlestick injuries occurred whilst performing an injection, however a concerning number occurred through bad practice (i.e. recapping a needles, passing the device, or sharps being in regular waste). Additionally, when active ingredients were mentioned, some included immunosuppressants and sedatives.

**Table 4:** Needlestick injury prevalence and mechanisms within companion animal practices.

|  | Recent Injuries (n=47) | Severe Injuries (n=22) |
| --- | --- | --- |
| Overall prevalence | 9.6% of all injuries | 4.8% of all injuries |
| Mechanism of needlestick |  |  |
| Injecting | 23.4% | 31.8% |
| Recapping | 21.3% | 31.8% |
| Fine Needle Aspirates | 8.5% | 9.1% |
| Blood Sample | 8.5% |  |
| Drawing Up | 8.5% |  |
| Intravenous Cannula Placement | 6.4% | 4.5% |
| Handling / Passing Device | 6.4% |  |
| Handling Waste (Sharps Bin/Bag) | 4.3% |  |
| Knocked over by animal | 2.1% | 4.5% |
| Unknown | 10.6% | 18.2% |
| Active ingredients mentioned: | Cyclosporine<br>Methadone<br>Pentobarbital<br>Propofol<br>Vaccines (Nobivac Tricat Trio, Nobivac L4) | Meloxicam<br>Methadone<br>Pentobarbital<br>Propofol |

### Context of animal-related injuries

In 21.4% of recent, and 18.2% of severe, animal-related injuries the injured person describes the animal ‘attacking out of nowhere’ or that it ‘suddenly bit’ or that there were no behavioural warning signs.

> “*Was trying to look at a wound and dog went for my arm with no warning*.” – VN, dog bite to arm, received first aid

In 9.3% of recent, and 9.5% of severe, animal-related injuries the injured person describes that the animal was fearful/anxious preceding the injury.

> “Dog was a bit nervous but the owner said she liked humans, just dogs she didn’t. As I just had palpated her abdomen and was raising myself up she launched at me and bit my face.” – VS, dog bite to face, received first aid

When another person was present at the point of injury, 2.7% of recent and 5.3% severe injury circumstances, the injured person blamed the other individual for the injury occurring.

> “*I was holding a German Shepherd Dog for a painful intramuscular injection, explained to Vet I didnt have a good hold on him but Vet went ahead and injected, dog then bit me on forearm*” – VN, dog bite to arm, received first aid.

Muzzling of an animal was only mentioned in 2.2% of recent animal-related injuries, however this rose to 5.7% when related to severe injuries. The common mechanism for muzzle related injuries were, in order of frequency; the dog bit through the muzzle, the dog bit when the muzzle was being putting on, the dog removed the muzzle, and that the dog was in post anaesthetic recovery when it was being muzzled.

> “*Bitten by an aggressive dog when we were trying to euthanise him. He had a muzzle on and had been given heavy sedatives but he completely freaked out, got the muzzle off and attached himself to my arm*” – VN, dog bite to arm, visited hospital emergency department

In a few circumstances the owner of the dog was asked to muzzle the dog, which they refused, and subsequently veterinary staff were injured.

> “*Vet wanted bloods done in consult in front of owner, wouldn’t allow muzzle to be put on, dog chewed on my finger”* – Vet, dog bite to arm, attended primary care physician

### Injury consequences and behavioural response to injury

Overall, 1.0% (95% CI 0.3-2.4) of companion animal practice staff’s most recent work-related injury was reportable to the HSE through RIDDOR as a ‘specified injury’. This did not vary between role: administrative 4.2% (95% CI 0.5-14.3), receptionist 2.9% (95% CI 0.1-14.9), ACA’s 2.3% (95% 0.1-12.3), and VS 0.5% (95% CI 0.0-3.0); VNs had no reportable injuries. However, when looking at their most severe injuries, 4.3% (95% CI 2.7-6.6) were RIDDOR reportable as a ‘specified injury.’ This did not vary between role: administrative 6.7% (95% CI 1.4-18.3), receptionist 3.2% (95% CI 0.1-16.7), ACA 2.6% (95% CI 0.1-13.5), VN 3.5% (95% CI 1.3-7.4), and VS 5.3% (95% CI 2.4-9.8).

As most animal-related injuries occurred in veterinarians and VNs and were related to cats and dogs, we focused on these groups to understand the anatomical distribution of injuries. Top cat-related injuries to VNs were bites to hands (recent: 48.4%, severe: 56.6%), scratch to hands (recent: 19.8%, severe: 10.8%), scratch to arms (recent: 13.2%, severe: 9.6%), and bites to arms (recent: 12.1%, severe: 10.8%). Whilst those to vets were bites to hands (recent: 62.9%, severe: 63.1%) scratch to hands (recent: 23.5%, severe: 8.7%), bite to arms (recent: 11.8%, severe 10.9%), and scratch to arms (recent: 11.8%, severe: 8.7%) (S2).

Top dog-related injuries to VNs involved bites to hands (recent: 38.3%, severe: 30.0%), bites to arms (recent: 26.8%, severe: 26.0%), scratches to arms (recent: 17.1%, severe: 2.0%), ergonomic injuries (recent: 4.9%, severe: 14.0%) and bite to heads (recent: 4.9%, severe: 6.0%). Whilst those to vets were bites to hands (recent: 47.0%, severe: 51.4%), bites to arms (recent: 20.4%, severe: 17.1%), and bites to the head (recent: 8.2%, severe: 12.8%). Two percent of severe VN dog-related injuries resulted in concussion, and a further 2% resulted in fracture. Four percent of veterinarians most recent dog-related injuries resulted in fractures; this was similar for the most severe injuries (4.2%). Whilst 5.7% of severe injuries were degloving. On exploration of the events preceding bites to the head, they all occurred whilst in the waiting or consult room and were associated with either the clinical examination of the head or the dog being passed in the owner’s arms to the VS or VN. Owners were always present for this type of injury.

Most staff were injured with someone else present (Table 5, S3). For all roles, excluding veterinarians, the main individuals present were either VNs or VS. In contrast for veterinarians, the main individuals present at injury were an animal’s owner or VN. The minority of injuries did not receive medical attention. The main form of medical attention for all roles was self-administered first aid. Dependent on role, between 16.3% (VN) and 23.0% (receptionists) of staff attended or were admitted to hospital for their most recent injury. Considering their most severe injuries this increased to between 25.8% (receptionists) and 36.9% (VN).

**Table 5:** Consequences of recent work-related injuries in companion animal veterinary practices †These sections won’t always add to 100% as multiple people could be present at an injury.

|  | Administrative staff (n=48)<br>(95% CI) | Reception Staff (n=35)<br>(95% CI) | Animal Care Assistants (n=40)<br>(95% CI) | Veterinary Nurses (n=179)<br>(95% CI) | Veterinary Surgeons (n=171)<br>(95% CI) |
| --- | --- | --- | --- | --- | --- |
| **Was anyone else present at point of injury? †** |  |  |  |  |  |
| No | 29.6% (17.0-44.1) | 23.5% (10.8-41.2) | 15.0% (5.7-29.8) | 19.6% (14.0-26.1) | 8.2% (4.5-13.4) |
| Admin | 2.1% (0.1-11.1) | 2.9% (0.1-14.9) |  | 1.1% (0.1-4.0) |  |
| Animal Care Assistant | 4.2% (0.5-14.3) |  | 5.0% (0.6-16.9) | 4.5% (1.9-8.6) | 1.2% (0.1-4.2) |
| Owner or client | 10.4% (3.5-22.7) | 11.4% (3.2-26.7) | 10.0% (2.8-23.7) | 5.0% (2.3-9.3) | 38.0% (30.7-45.7) |
| Receptionist | 6.3% (1.3-17.2) | 8.8% (1.9-23.7) | 2.5% (0.1-13.2) | 2.2% (0.6-5.6) | 1.2% (0.1-4.2) |
| Student Vet Nurse |  |  | 5.0% (0.6-16.9) | 4.5% (1.9-8.6) | 2.3% (0.6-5.9) |
| Vet Nurse | 22.9% (12.0-37.3) | 29.4% (15.1-47.5) | 30.0% (16.6-46.5) | 26.8% (20.5-33.9) | 44.4% (36.9-52.2) |
| Vet Student | 4.2% (0.5-14.3) | 2.9% (0.1-14.9) |  | 0.6% (0.0-3.1) |  |
| Vet Surgeon | 20.8% (10.5-35.0) | 14.3% (4.8-30.3) | 47.5% (31.5-63.9) | 44.7% (37.3-52.3) | 8.8% (5.0-14.1) |
| Other |  |  |  |  |  |
| **Medical Treatment** |  |  |  | 178 |  |
| None | 8.3% (2.3-20.0) | 14.3% (4.8-30.3) | 5.0% (0.6-16.9) | 7.3% (3.9-12.2) | 21.6% (15.7-28.6) |
| Self-administered first aid | 58.3% (43.2-72.4) | 34.3% (19.1-52.2) | 52.5% (36.1-68.5) | 55.6% (48.0-63.1) | 46.2% (38.6-54.0) |
| First aid received | 6.3% (1.3-17.2) | 22.9% (10.4-40.1) | 10.0% (2.8-23.7) | 2.8% (0.9-6.4) | 2.9% (1.0-6.7) |
| Primary Care | 4.2% (0.5-14.3) | 5.7% (0.7-19.2) | 10.0% (2.8-23.7) | 17.4% (12.2-23.8) | 9.9% (5.9-15.4) |
| Minor Injury Unit | 8.3% (2.3-20.0) | 14.3% (4.8-30.3) | 17.5% (7.3-32.8) | 12.4% (7.9-18.1) | 12.9% (8.2-18.8) |
| Emergency Department | 10.4% (3.5-22.7) | 2.9% (0.1-14.9) | 2.5% (0.1-13.2) | 2.2% (0.6-5.7) | 5.3% (2.4-9.7) |
| Hospital Admission | 4.2% (0.5-14.3) | 2.9% (0.1-14.9) | 2.5% (0.1-13.2) | 1.7% (0.3-4.8) | 1.2% (0.1-4.2) |
| Hospital Outpatients |  | 2.9% (0.1-14.9) |  |  |  |
| Physiotherapist |  |  |  | 0.6 (0.0-3.1) |  |
| **Physical Recovery Time** |  |  |  |  |  |
| < 7 days | 66.7% (51.6-79.6) | 65.7% (47.8-80.9) | 75.0% (58.8-87.3) | 65.7% (58.3-72.7) | 74.9% (67.7-81.2) |
| 7-14 days | 8.3% (2.3-20.0) | 20.0% (8.4-36.9) | 17.5% (7.3-32.8) | 27.0% (20.6-34.1) | 18.1% (12.7-24.7) |
| 15-28 days | 12.5% (4.7-25.3) |  | 5.0% (0.6-16.9) | 3.9% (1.6-7.9) | 4.1% (1.7-8.3) |
| >28 days | 12.5% (4.7-25.3) | 14.3% (4.8-30.3) | 2.5% (0.1-13.2) | 3.4% (1.2-7.2) | 2.9% (1.0-6.7) |
| **Time off work** |  |  |  |  |  |
| None | 85.4% (72.2-93.9) | 88.6% (73.3-96.8) | 85.0% (70.2-94.3) | 90.4% (85.2-94.3) | 91.8% (86.7-95.5) |
| <7 days | 4.2% (0.5-14.3) |  | 12.5% (4.2-26.8) | 6.7% (3.5-11.5) | 7.0% (3.7-11.9) |
| 7-14 days | 2.1% (0.1-11.1) | 2.9% (0.1-14.9) |  | 1.1% (0.1-4.0) | 0.6% (0.0-3.2) |
| 15-28 days | 2.1% (0.1-11.1) | 2.9% (0.1-14.9) |  |  | 0.6% (0.0-3.2) |
| >28 days | 6.3% (1.3-17.2) | 5.7% (0.7-19.2) | 2.5% (0.1-13.2) | 0.6 (0.0-3.1) |  |
| **Was there a practice policy to minimise risk?** |  |  |  |  |  |
| Yes | 41.7% (27.6-56.8) | 11.4% (3.2-26.7) | 30.0% (16.6-46.5) | 15.2% (10.2-21.3) | 8.8% (5.0-14.1) |
| No/Not aware | 58.3% (43.2-72.4) | 88.6% (73.3-96.8) | 70.0% (53.5-83.4) | 84.8% (78.7-89.8) | 91.2% (85.9-95.0) |
| **Did you inform practice management?** | 47 | 34 |  |  |  |
| Yes | 85.1% (71.7-93.8) | 76.5% (58.8-89.3) | 97.5% (86.8-99.9) | 87.6% (81.9-92.1) | 69.0% (61.5-75.8) |
| **Did you file an accident/incident report?** |  |  |  |  |  |
| Yes | 61.7% (46.4-75.5) | 70.6% (52.5-84.9) | 90.0% (76.3-97.2) | 74.1% (67.1-80.4) | 47.4% (39.7-55.1) |
| **Was any subsequent action taken by the practice?** |  | 32 |  | 176 |  |
| Yes | 23.4% (12.3-38.0) | 15.6% (5.3-32.8) | 15.0% (5.7-29.8) | 18.8% (13.3-25.3) | 5.3% (2.4-9.8) |
| No | 66.0% (50.7-79.1) | 59.4% (40.7-76.3) | 65.0% (48.3-79.4) | 69.9% (62.5-76.6) | 78.4% (71.4-84.3) |
| Don't know | 8.5% (2.4-20.4) | 25.0% (11.5-43.4) | 20.0% (9.1-35.7) | 18.8% (13.3-25.3) | 16.4% (11.2-22.8) |

The majority of staff did not experience any mental or emotional effects resultant of the injury (Table 6, S4). (Further supportive quotes to this section of results can be found in Supplementary Material 4 when indicated.) When they did the main effect was ‘increased anxiety or fear’ when in the same situation (S4).

**Table 6:** Reasons for behavioural responses to a recent work-related injury in companion animal veterinary practices.

| Themes | Administrators | Receptionists | Animal Care Assistants | Veterinary Nurses | Veterinary Surgeons |
| --- | --- | --- | --- | --- | --- |
| Did you experience any **emotional or mental** effects resultant of the injury? | n=47 | n=35 | n=40 | n=178 | n=171 |
| None | 91.5% | 91.4% | 90.0% | 87.6% | 83.6% |
| Increased anxiety or fear |  | 5.7% | 10.0% | 6.7% | 12.3% |
| More cautious around the animal |  |  |  | 3.9% | 4.1% |
| Won't work with that species again | 2.1% |  |  | 0.6% |  |
| Confidence knocked |  |  |  | 0.6% | 0.6% |
| Embarrassment | 2.1% | 2.9% |  | 0.6% | 1.2% |
| Annoyance | 2.1% |  |  | 0.6% |  |
| Undervalued by employer | 2.1% |  |  | 1.1% | 0.6% |
| Why did you **not take time off** from work? | n=40 | n=31 | n=34 | n=161 | n=157 |
| Genuinely minor injury not requiring time off work | 85.0% | 93.5% | 97.1% | 89.4% | 85.4% |
| Could still function with a reduced workload | 10.0% | 6.5% |  | 8.1% | 8.9% |
| 'Be tough'/'Just get on with it!' |  |  |  | 1.9% | 1.3% |
| Don't want to lose sick leave/sick pay etc | 2.5% |  |  | 0.6% |  |
| Not allowed to by employer |  |  |  |  | 1.3% |
| Used annual leave | 2.5% |  | 2.9% | 1.2% | 2.5% |
| Didn't want to let team down |  |  |  | 0.6% | 3.2% |
| Why did you choose **not to report** the incident? | n=18 | n=8 | n=4 | n=45 | n=90 |
| Only minor injury, which wasn't serious enough | 22.2% | 75.0% | 50.0% | 64.4% | 54.4% |
| Unaware it needed reporting |  |  |  |  | 3.3% |
| Didn't want to make a fuss |  |  |  | 2.2% | 2.2% |
| Too much effort as was too busy | 16.6% | 12.5% |  | 15.6% | 16.7% |
| It would cause tension amongst the team |  |  |  | 4.4% |  |
| It's just one of those things/'par for the course' | 5.6% |  | 25.0% | 8.9% | 8.9% |
| Not employed at CVS at the time | 38.9% | 12.5% | 25.0% | 2.2% | 2.2% |
| I forgot to do it | 27.8% |  |  | 13.3% | 12.2% |
| No reporting system in place | 5.6% |  |  | 2.2% | 11.1% |
| Have you **changed your behaviour** around the animal species involved in the injury? | n=44 | n=34 | n=39 | n=176 | n=169 |
| No | 59.1% | 47.1% | 48.7% | 47.2% | 52.1% |
| More cautious and less confident | 6.8% | 2.9% | 7.7% | 11.4% | 9.5% |
| Avoidance of procedure or animal | 2.3% |  | 2.6% | 0.6% | 2.4% |
| Do procedure differently | 6.8% | 14.7% | 15.4% | 14.8% | 10.1% |
| Improve handling/restraint | 6.8% | 2.9% | 15.4% | 10.8% | 9.5% |
| Would now use personal protective equipment or a muzzle | 11.4% | 2.9% | 7.7% | 9.1% | 11.2% |
| Denial that did anything wrong | 2.3% |  |  | 3.4% | 3.0% |
| Improved awareness of surroundings | 9.1% | 14.7% | 5.1% | 5.1% | 4.7% |

> “*It was a real shock as I had previously felt fairly confident with dogs and knowing if they were at risk of showing aggression. I am now pretty fearful of German Shepherds in particular! The fact the dog went for my throat was what scared me the most - although the hand injury wasn’t too severe, it could have been severe or impacted my face*.” – VN, dog bite to hand, attended hospital emergency department

However, a range of effects were mentioned including feeling ‘undervalued by the employer’, being ‘more cautious around that species of animal’, ‘embarrassment’ and having their ‘confidence knocked’.

More than a quarter of all roles felt they needed more than seven days to physically recover from their most recent injury (Table 5). This increased when the injury was more severe, most notably in VN and VS in which 64.2% and 57.3% respectively needed more than seven days. However, there was discordance between time to recover and time taken off work. Overall, less than 14% of injured persons took more than seven days of work. This discordance was most noted in severe injuries to veterinarians, in which <5% took more than seven days off work. This was despite almost 10% of the injuries involving fractures or degloving’s. The main reason the injured party did not want to take time off was that the injury was genuinely minor, and it did not require time off work (Table 6). Overall, this reason accounted for more than 85% of the responses. Other reasons included that they ‘did not want to let the team down’:

‘*I could still drive and was physically able to work, and didn’t want my colleagues to have to do my work for me*.’ – VS, dog bite to face, attended minor injuries unit.

In some circumstances they felt that they could still work, but with a reduced workload (S4), a few gave the impression that they needed to “be tough” and “get on with it” (S4):

> “*I can fight through it, it is imperative that I work*” – VS, manual handling injury, attendance at hospital emergency department

In a minority of circumstances, the injured persons manager would not let them take time off work:

> “*I asked to but was told my practice would give me light duties. They didn’t*.” – VS, chronic shoulder tendonitis, attendance at primary care physician

In all roles, except veterinarians, the majority of respondents reported their injury via an official accident book or injury reporting system. Most veterinarians did not report their injuries. The main three reasons given were; It was ‘too much effort’ or they were ‘too busy’ to fill in the report (S4);

> “*I did not have time for paperwork and I could manage it myself I didn’t need people checking up on me*” – VS, cat bite to hand, received first aid

Some veterinarians saw that their injury was inevitable and just an everyday hazard not worthy of reporting and some just forgot to report it (S4);

> “*Part of the job. Plus far too many questions and not enough time to complete, then get chased for more info I don’t have time to answer*” – VS, dog bite to hand, attendance at hospital emergency department
>
> “*It was less of conscious decision and more due to the fact that it was a busy day with more pressing matters to attend to and by the next day reporting the incident was forgotten about*.” – VS, dog bite to hand, received first aid

In a minority of cases, respondents did not want to be perceived differently by their colleagues;

> “*I was much less confident about reporting and didn’t want to make a fuss or seem high maintenance therefore I got on with it*” – VS, cat bite to hand, no treatment

Around half of respondents would change their behaviour if placed in the same scenario, for their most recent injuries, this increased for all roles when the injury was their most severe. This concerningly still meant a high proportion would not change their behaviour, and thus place themselves at risk again. Some individuals would avoid the procedure or animal to totally negate the risk.

> “*I try to avoid any contact with fractious cats if I can*.*”* – VS, cat bite to hand, attendance at hospital emergency department
>
> “*I’m no longer allowed to handle squirrels*” – ACA, squirrel bites to hand and arms, attendance at hospital emergency department

The change in behaviour was split between four key themes. Firstly, that the respondent was ‘more cautious, and less confident’ in the same situation (S4);

> “*I’ll just be more careful when resheathing needles*” – VN, needlestick to hand, received first aid

Some respondents would carry out the procedure differently (S4);

> “*I read a lot and learnt a lot about dog behaviour so I am much more aware of body language and I would never put myself in a similar position again*” – VS, dog bite to hand, received first aid

Many would improve their restraint techniques (S4);

> “*Now will try to towel wrap and also dispense ‘chill protocol’ prior to appointment*.” – VN, cat scratch to arm, received first aid

Finally, many would now start muzzling dogs on a more regular basis prior to consultations, examinations or procedures (S4);

> “*Muzzle more often and insist on them despite owner protests of ‘he doesn’t bite’, as this is followed by ‘he hasn’t done that before*’” – VS, dog bite to head, attendance at minor injury unit

Most respondents were not aware of, or did not think there was, any practice policy in place that could have mitigated their injury. Policies when mentioned included; cat and dog handling guidelines, feral cat and wildlife handling guidelines, and general personal protective equipment (PPE) guidelines. Similarly, when asked whether the practice, where they were employed at the time, took any action after the incident described, the majority said that no action was taken, or they were not aware of any action. When actions were described they included; a review of current risk assessments, a review of the incident, workload and rota adjusted to accommodate injured person, new practice muzzle policy, new sedated dogs protocol, review of chemical dilutions protocols, review of cleaning protocol, provision of animal handling training for all staff, and new PPE was provided.

## Discussion

This is one of the largest studies to explore work-related injuries within companion animal veterinary practices, and the first to explore all job roles. Roles primarily involving animals had a higher annual prevalence of injury (Mills 2019), a higher frequency of injuries (Johnson & Fritschi 2024; Tulloch *et al*. 2025), and were predominately injured by animals (Epp & Waldner 2012; Fritschi *et al*. 2006; Gabel & Gerberich 2002; Johnson & Fritschi 2024; Nienhaus *et al*. 2005; Tulloch *et al*. 2023). Thus, showing that animals are the main driver of injuries for veterinarians, nurses and ACAs. This is emphasised by our model where job role was the main significant variable associated with receiving an injury. This model showed that hours-worked was an important variable, with roles undertaking <20 hours or >40 hours of work a week at a generally lower risk to obtaining injuries compared to roles working 21 – 40 hours a week, however this relationship was non-linear and challenging to interpret. Further research is needed to understand this relationship, though it could be linked to differences in experience, animal exposure, or task allocation.

Around half of injuries to receptionists and administrators involved animals, primarily restraint of the animal. This highlights the multi-faceted nature of these roles, and that within a companion animal clinical setting, duties may not be fully clerical in nature. It is important that those in these roles, without prior veterinary experience, have been trained appropriately in how to handle and restrain animals. The remainder of injuries match similar national workplace injury trends with slips, trips, and strikes by equipment or furniture being the most common cause of injury (Jabbour *et al*. 2015; Lundstrom *et al*. 2023). Standard injury prevention guidance should be followed to minimise the risk of these injuries (Health & Safety Executive 2012).

The practice location of injuries (prep room, consulting room, ward) were reflective of where ACAs, VNs, and VS spend much of their time. This indicates that any interventions developed to minimise work-related injuries for clinical staff should be focused in these areas. It is unsurprising that most injuries were animal-related with the main species involved being cats and dogs (Johnson & Fritschi 2024; Tulloch *et al*. 2023). Restraint of cats appeared to be a particular issue especially when they were being held by VNs and ACAs for blood samples, injections, and clinical examinations. Cat friendly veterinary interaction guidelines do exist (Rodan *et al*. 2022), and an estimated 37% of UK practices (based on survey responses) are designated cat friendly clinics (Feilberg *et al*. 2021). Restraining cats can be challenging and unpredictable, yet these results indicate that potentially this guidance and training may not be working or adhered to in these situations, and that more training is needed across the profession to improve cat handling and restraint. During these restraint procedures, it was notable that some injured persons blamed the other person involved in the restraint or veterinary procedure for the injury. This often led to a lack in trust of the ‘blamed’ individual. This potentially evidences team miscommunication and co-ordination, and differing expectations of their colleagues’ anticipated movements during the restraint process. Since some respondents attributed their injury on a colleague’s actions, it is possible that they do not see the need to learn or adapt their subsequent behaviour as it was out of their own personal control. A blame culture can harm team dynamics and lead to workplace tension. Shifting towards a learning culture could minimise injuries and improve communication. This would involve using a team-based approach where anyone involved in restraint had clearly defined roles and responsibilities. Inclusive reflection post injury, or near miss incident, would facilitate learning opportunities to improve restraint and could enable discussions about what could be done differently rather than attributing blame. Practices should foster a work environment where safety concerns can be raised openly and constructively without the potential fear for punishment.

VNs and VS were frequently injured when animals were recovering from anaesthesia, with the animal ‘suddenly waking up.’ This is similar to an audit of UK veterinary schools where they found that 64% of all head injuries caused by dogs were associated with dogs anaesthetic recovery or extubation (Tulloch *et al*. 2023). These results raise questions about the recovery environment, including anaesthetic depth, quality of pain relief, recognition of signs of disorientation, environmental calmness, and monitoring levels. Anaesthesia and monitoring guidelines do exist (Grubb *et al*. 2020), however they primarily focus on patient safety and do not discuss the potential of injury to veterinary professionals nor how they can be reduced. Further research is needed to understand what the most appropriate preventative measures are, and how they can balance patient safety and occupational safety.

Clinical examinations were the most common activity occurring at the point of injury for VS. Examinations to the head represented around half of these. These are likely due to defensive reactions, pain sensitivity, and restricted vision of the animal. Proper restraint, gentle handling, and a calm approach could help minimise risk. Understanding animal behaviour is key to reducing animal-related injuries during a consultation. One in five recent injuries were described as ‘out of nowhere’ attacks, where the IP reported no behavioural warning signs. In other incidents fear or anxiousness of the animal was seen, but no adjustment behaviour by the IP was subsequently described to reduce the likelihood of a negative physical interaction. Dogs and cats display complex and subtle emotional signals, that may not be overtly visible during a consultation or be difficult to interpret (Demirbas *et al*. 2016; Ellis 2018; Walsh *et al*. 2024). If these signs were missed, then it could appear that the animal attacked ‘out of nowhere.’ To reduce injuries from animals that are in fear or pain, or are aggressive, then a consultation environment needs to be created which enables low-stress handling techniques and reduces miscommunication between humans and animals. This could be facilitated through further animal behaviour training of those with roles who interact with animals, alongside interventions that could minimise the risk of injury, such as using muzzles. If muzzling or other PPE, restraint mechanisms are used, then complementary education of owners is required so that an appreciation for the muzzles need is acquired. Practice policy should be developed that supports VS and VNs who request a muzzle for a patient, but where an owner declines. Additionally, muzzles should be used more generally with appropriate sizes and fits for all types of dogs. As seen in our data, dogs can bite through a muzzle if it is incorrectly fitted.

The high prevalence of needlesticks is a concern; almost 10% of all injuries and 1 in 6 of VS injuries. This is lower than reported in Australian VSs (57%) and VNs (75%) (Johnson & Fritschi 2024). This could be explained by safety culture in the UK veterinary profession, also seen in Canada (Weese & Jack 2008). The veterinary professionals in our study have previously described needlestick injuries as a common incident not worthy of reporting (Furtado *et al*. 2024). They also describe needlesticks not fulfilling their criteria of an injury, namely that they were common, minor, self-attributed and not causing lasting pain. As such, it is likely that these prevalence figures represent a large under-reporting of the true prevalence. In our results, a third of these injuries were through not following recommended guidance (i.e. recapping of needles, passing the device, or sharps being placed in regular waste). These injuries pose a risk to IPs through exposure to zoonotic infections (ie *Brucella canis*) and potential exposure to bloodborne pathogens (ie hepatitis) (Robertson *et al*. 2016). More common place is that they can lead to injection site reactions, abscess formation, or secondary ascending infections if bacterial pathogens enter the wound (Robertson *et al*. 2016). Accidental injections of veterinary drugs could cause serious health effects (including allergic reactions or systemic toxicity), and many of the drugs discussed in our results were immunosuppressants and sedatives. Needlestick injuries, while common in veterinary practice, should always be taken seriously to prevent long-term health risks. More research is needed to understand the veterinary professions attitudes towards needlesticks injuries, to best develop appropriate workplace safety improvements.

Only a small proportion of injuries experienced were a RIDDOR reportable injury, in contrast to equine and production animal vets where 11.6% and 13.3% of their respective injuries were reportable (Tulloch *et al*. 2025). Cat-related injuries were predominately bites to hands, and scratches to hands and arms, consistent with previous literature (Colmers-Gray *et al*. 2023; Fritschi *et al*. 2006; Kheiran *et al*. 2019; Tulloch *et al*. 2023). Twenty-eight percent of cat bite injuries led to attendance or admission at a hospital, this is much higher than the 5% reported at UK veterinary schools (Tulloch *et al*. 2023). Current recommendations are that most cat bites should receive antibiotic prophylaxis, due to their high risk of infection (Colmers-Gray *et al*. 2023). These figures represent an improvement from previous literature, but further education of the veterinary community is needed to encourage rapid assessment of cat bite injuries at emergency departments. Dog-related injuries were mainly bites to hands and arms, as per the literature (Colmers-Gray *et al*. 2023; Fritschi *et al*. 2006; Tulloch *et al*. 2023, 2021) however almost one in ten were bites to heads. Previous research had found that bites to the head were associated with the animal’s anaesthetic recovery (Tulloch *et al*. 2023), where as we found that they were associated with clinical examinations of the head and the passing of the dog between the owner and IP. These are both common daily activities in veterinary practice and investigations need to be performed to assess the best methodology to minimise the injury risk from them.

VNs experienced ergonomic injuries, mostly through lifting heaving patients or equipment. Research from Australia has identified that more than half of VNs suffered from chronic back or neck pain (van Soest & Fritschi 2004), most likely from poor ergonomic practices. VNs do a lot of heavy lifting of patients, with larger dog breeds being in excess of 50kg. Lifting heavy patients increases muscular and joint pain, compared to a rigid object, as the dog can unpredictably move or be limp if sedated/unconsciousness. Practices should encourage team lifting for large dogs and adopt appropriate lifting techniques. The development and utilisation of mechanical lifting aids should be explored to reduce the need form manual lifting and reduce the burden of musculoskeletal injuries on practices.

More than 1 in 10 injuries to VS were lead-related. These types of injuries can be serious, noted by the fractures to feet and hands seen in our data. The common mechanism for these injuries tend to result from a pull with a subsequent trip or tangle in the lead (Forrester 2020). These injuries are likely due to a combination of factors including inappropriate lead restraint (i.e. extendable or long leads) and reduced situational awareness. These should represent some of the more preventable injuries within veterinary practices. Solutions could include using team-based handling for larger or agitated animals, and provision of appropriate restraint tools if the dog’s owner does not provide appropriate restraint.

Most individuals did not experience emotional or mental health consequences resultant of the injury. Studies have shown that traumatic workplace injuries are associated with worse mental health trajectories than non-workplace injuries (Wightman *et al*. 2025), and that individuals injured at work have worse self-reported general health and mental health ten years post-injury (Dong *et al*. 2015). Considering that the veterinary profession has a high prevalence of mental health issues (predominately anxiety and depression) and suicide (Bartram *et al*. 2009; Platt *et al*. 2010), it is surprising that most respondents were not impacted by their injuries, especially considering the severity of some of the injuries. Possible reasons for a low prevalence of mental health issues could include denial, unaware of mental health symptoms or an inability to recognise them, cultural stigma, or masking through strong coping mechanisms. Research is needed to understand this issue.

The most common form of medical treatment was self-administered first aid. However, there was a higher reported attendance at hospital for VNs (16.3%) and VSs (19.4%), than in previous UK literature (5% of cat-related, and 4% of dog related injuries to clinical staff at veterinary schools) (Tulloch *et al*. 2023). For comparison this was 25.1% for equine veterinarians, and 40.0% for production animal veterinarians, in general practice (Tulloch *et al*. 2025). These higher rates of hospital attendance could have multiple potential causes that require further exploration to understand. There could be self-selection bias, as those who have experienced more severe injuries may have been more inclined to participate in the study. There may be more stringent health and safety standards at the participant’s veterinary practice where the injury occurred, whereby the threshold for hospital attendance is lower than other organisations. There could be more awareness and reduced stigma around injury and so that injuries are being taken seriously, or that these respondents are receiving more serious injuries than the prior literature.

Similar to previous research there was discordance with the time that an IP took to physically recover, and the time taken off work (Tulloch *et al*. 2025). For most individuals the injury was minor enough that an IP potentially would have not needed to take time off work. Some felt that they could function with a lightened or adapted workload. In comparison to equine veterinarians there was much less bravado and dismissive language around severe injuries, and a lower percentage of people feeling guilty about time off work or letting their colleagues down (Tulloch *et al*. 2025). This potentially indicates that different safety cultures exist between companion animal and large animal vets, and that any interventions developed to change these cultures should be tailored to each broad veterinary sector. The veterinary industry as a whole should be aware that working whilst injured increases the risk of re-injury significantly in the first 6 months after injury, but can impact the IP’s health for up to four years (Sears *et al*. 2021). Our results identify a strong culture of presenteeism, the consequences of which in other industries are well described. Individuals will have decreased mental and physical acuity and health, with reduced productivity (Sanderson & Cocker 2013; Skagen & Collins 2016). Whilst the practice will have a decline in practice culture, patient care, staff wellbeing, reduced team cohesion, and increases in accidents and job turnover (Sanderson & Cocker 2013). This culture needs to be explored further in the veterinary industry with industry-specific interventions developed.

Around half of respondent would not alter their behaviour if placed in a similar situation to the one that led to their injury. This is proportion is notably higher than that reported in equine and production animal clinical staff (Tulloch *et al*. 2025). However, respondents who experienced more severe injuries were more likely to report subsequent behavioural changes. When changes were made these typically involved modifying procedural technique, exercising greater caution cautious, and utilising better restraint or PPE. These findings show the need for fostering a stronger culture of shared responsibility and reflective practice within veterinary teams. Structured debriefs and team-based safety training may support this shift, ensuring that all staff feel both empowered and accountable for workplace safety.

The awareness and knowledge of practice health and safety protocols and policies were poor, with the majority not knowing of any policy. This partnered with most VSs not reporting injuries, is indicative of a work safety culture where formal procedures could be poorly communicated or under-valued. This could contribute to under-reporting, lack of accountability and missed opportunities for learning and prevention. This is exemplified by the minimisation of reporting by VSs where they general felt that reporting took too much effort and that workplace injuries were par for the course. This dismissive tone, often carrying a ‘badge of honour’ attitude, is problematic as it undermines safe working practices and reflects a misunderstanding of the importance of injury reporting for the health and wellbeing of colleagues. This is similar to large animal clinicians who normalise workplace injuries as inevitable (Tulloch *et al*. 2025).

This work has some limitations. By making all veterinary roles eligible to respond, we limited our ability to explore role specific factors and behaviours in depth. Response rates may have improved if we had disseminated role specific questionnaires. Future studies on work-related injuries should focus both on specific roles and the interaction between roles. While a substantial number of individuals completed the survey, some did drop out; 25 VNs, 10 administrators, and 7 VS. Anticipating this, we offered an incentive available only upon survey completion. We made clear that we were asking respondents about injuries that may or may not have occurred in CVS practices, their current employers. However, a few participants noted that they did not report the injury in the survey as they were not at CVS at the time. We are also conscious that some individuals may have given responses of what they felt their managers would prefer to hear. However, we believe that this bias was minimal due to the anonymised nature of the survey, and that only summary data was shared with CVS. This is evidenced by the openness and honesty of numerous free-text responses.

## Conclusions

This study highlights the high prevalence of work-related injuries that occur within companion animal practices. We have identified key areas where injuries are common and where research is needed to identify best practices or interventions that can help minimise risk and prevent injuries. These areas include; cat handling, general animal restraint, needlestick injuries, and animals recovering from anaesthetics. A culture of presenteeism and blame appears to be embedded in these settings, where the personal and institutional impact of injuries are routinely minimised at the potential detriment of all. A health and safety cultural shift is needed; one that moves from blame to shared responsibility and reflective learning. Achieving this will require strong organisational leadership and a commitment to fostering an environment where all staff feel they speak openly and take accountability for workplace safety.

## Data Availability

All data produced in the present study are available upon reasonable request to the authors

## Conflict of Interest

IS and RJ are current employees of CVS UK Ltd.

## Acknowledgements

The authors would like to thank all the participants of this study for their frank and honest viewpoints.

## Author Contributions

Conceptualisation: JT. Funding Acquisition – JT, MW, RJ. Methodology: JT, MW, IS. Formal analysis: JT. Writing original draft – JT. Writing review & editing – All authors

## Funding

This work was funded by the CVS Clinical Research Awards (PRA00009, 2022).

## Notes

### Author Declarations

The study received ethical approval from the University of Liverpool Veterinary Research Ethics Committee (VREC1256).

