## Supplementary Material for "‘Just part of the job’ – understanding work-related injuries and safety culture in companion animal veterinary practices"

**S1 Context of severe veterinary work-related injuries in companion animal practices.** †Will be lower than animal involvement in injury. For example, sharps are not classified as having animal involvement, but if occurred during surgery an individual may say an animal was involved.

| | **Administrative staff (n=45) (95% CI)** | **Reception Staff (n=32) (95% CI)** | **Animal Care Assistants (n=39) (95% CI)** | **Veterinary Nurses (n=174) (95% CI)** | **Veterinary Surgeons (n=171) (95% CI)** |
|---|---|---|---|---|---|
| **When did the injury occur?** | | | | | |
| <3 months ago | 2.2% (0.1-11.8) | 9.4% (2.0-25.0) | 28.2% (15.0-44.9) | 8.6% (4.9-13.8) | 7.0% (3.7-11.9) |
| 3-6 months ago | | 9.4% (2.0-25.0) | 25.6% (13.0-42.1) | 6.3% (3.2-11.0) | 8.2% (4.5-13.4) |
| 7-9 months ago | 2.2% (0.1-11.8) | 3.1% (0.1-16.2) | 7.7% (1.6-20.9) | 9.2% (5.3-14.5) | 3.5% (1.3-7.5) |
| 10-12 months ago | 2.2% (0.1-11.8) | 15.6% (5.3-32.8) | 12.8% (4.3-27.4) | 6.3% (3.2-11.0) | 8.2% (4.5-13.4) |
| >12 months ago | 93.3% (81.7-98.6) | 62.5% (43.7-78.9) | 25.6% (13.0-42.1) | 69.5% (62.1-76.3) | 73.1% (65.8-79.6) |
| **Location of Injury** | | | | n=173 | |
| **Practice** | **95.6% (84.9-99.5)** | **96.9% (83.8-99.9)** | **97.4% (86.5-99.9)** | **98.3% (95.0-99.6)** | **82.5% (75.9-87.8)** |
| Bathroom | | | | | |
| Car Park | 2.2% (0.1-11.8) | 6.3% (0.8-20.8) | 2.6% (0.1-13.5) | 1.7% (0.4-5.0) | 2.3% (0.6-5.9) |
| Consult/Examination room | 8.9% (2.5-21.2) | 18.8% (7.2-36.4) | 17.9% (7.5-33.5) | 14.5% (9.6-20.6) | 36.8% (29.6-44.5) |
| Corridor | | 9.4% (2.0-25.0) | | 3.5% (1.3-7.4) | 0.6% (0.0-3.2) |
| Dispensary | | | 2.6% (0.1-13.5) | 0.6% (0.0-3.2) | 0.6% (0.0-3.2) |
| Isolation | | | 2.6% (0.1-13.5) | 1.7% (0.4-5.0) | |
| Kitchen | | 9.4% (2.0-25.0) | | | 0.6% (0.0-3.2) |
| Laboratory | | | | 0.6% (0.0-3.2) | 0.6% (0.0-3.2) |
| Laundry | 2.2% (0.1-11.8) | | | | |
| Office | 2.2% (0.1-11.8) | 3.1% (0.1-16.2) | | | |
| Outside | 13.3% (5.1-26.8) | | 2.6% (0.1-13.5) | 2.9% (0.9-6.6) | |
| Prep room | 31.1% (18.2-46.7) | 9.4% (2.0-25.0) | 43.6% (27.8-60.4) | 43.4% (35.9-51.1) | 26.3% (19.9-33.6) |
| Radiography | | | 5.1% (0.6-17.3) | 2.3% (0.6-5.8) | 2.3% (0.6-5.9) |
| Reception | 8.9% (2.5-21.2) | 25.0% (11.5-43.4) | | 0.6% (0.0-3.2) | 1.2% (0.1-4.2) |
| Staff room | | | | | 0.6% (0.0-3.2) |
| Theatre | | | 2.6% (0.1-13.5) | 5.8% (2.8-10.4) | 3.5% (1.3-7.5) |
| Utility room | 6.7% (1.4-18.3) | | 2.6% (0.1-13.5) | | |
| Ward/Kennels | 20.0% (9.6-34.6) | 12.5% (3.5-29.0) | 15.4% (5.9-30.5) | 20.2% (14.5-27.0) | 5.8% (2.8-10.5) |
| Unknown | | | | 1.2% (0.1-4.1) | 1.8% (0.4-5.0) |
| **Non-Practice Location** | **4.4% (0.5-15.2)** | **3.1% (0.1-16.2)** | **2.6% (0.1-13.5)** | **1.7% (0.4-5.0)** | **17.5% (12.2-24.1)** |
| Animal Shelter | | | | | 0.6% (0.0-3.2) |
| Educational Institutes | 2.2% (0.1-11.8) | 3.1% (0.1-16.2) | | | 0.6% (0.0-3.2) |
| Farm | | | | 0.6% (0.0-3.2) | 7.0% (3.7-11.9) |
| Home visit | | | 2.6% (0.1-13.5) | | 1.8% (0.4-5.0) |
| Stable | | | | 0.6% (0.0-3.2) | 6.4% (3.3-11.2) |
| Zoo | 2.2% (0.1-11.8) | | | | |
| Unknown | | | | 0.6% (0.0-3.2) | 0.6% (0.0-3.2) |
| **Animal involvement in injury** | | | | | |
| None | 44.4% (29.6-60.0) | 53.1% (34.7-70.9) | 15.4% (5.9-30.5) | 16.2% (11.0-22.5) | 12.2% (7.8-18.2) |
| Cat | 20.0% (9.6-34.6) | 18.8% (7.2-36.4) | 48.7% (32.4-65.2) | 48.0% (40.3-55.7) | 28.7% (22.0-36.1) |
| Chicken | | | 2.6% (0.1-13.5) | | |

| | | | | | |
|---|---|---|---|---|---|
| Cow | | | | | 6.4% (3.3-11.2) |
| Degu | | | | 0.6% (0.0-3.2) | |
| Dog | 26.7% (14.6-41.9) | 25.0% (11.5-43.4) | 25.6% (13.0-42.1) | 32.9% (26.0-40.5) | 42.7% (35.2-50.5) |
| Ferret | 2.2% (0.1-11.8) | | | 0.6% (0.0-3.2) | 0.6% (0.0-3.2) |
| Gerbil | | | | | 0.6% (0.0-3.2) |
| Hamster | | | | | 0.6% (0.0-3.2) |
| Horse | 2.2% (0.1-11.8) | | | 1.2% (0.1-4.1) | 6.4% (3.3-11.2) |
| Parrot | 2.2% (0.1-11.8) | | | | |
| Pig | | | | | 0.6% (0.0-3.2) |
| Primate | | | 2.6% (0.1-13.5) | | |
| Rabbit | | | | | 0.6% (0.0-3.2) |
| Rat | | 3.1% (0.1-16.2) | | | |
| Raptor | | | 2.6% (0.1-13.5) | | |
| Squirrel | | | 2.6% (0.1-13.5) | 0.6% (0.0-3.2) | 0.6% (0.0-3.2) |
| Unknown | 2.2% (0.1-11.8) | | | | |
| **Mechanism of Injury** | | N=31 | | | |
| **Animal Injured Person †** | **51.1% (35.8-66.3)** | **48.4% (30.2-66.9)** | **74.4% (57.8-87.0)** | **75.1% (68.0-81.4)** | **81.9% (75.3-87.3)** |
| Administering oral medications | 2.2% (0.1-11.8) | | | | 1.8% (0.4-5.0) |
| Anaesthesia/induction/recovery | | 3.2% (0.1-16.7) | 2.6% (0.1-13.5) | 9.8% (5.8-15.3) | 5.8% (2.8-10.5) |
| Blood sampling | 2.2% (0.1-11.8) | | 2.6% (0.1-13.5) | 1.2% (0.1-4.1) | 1.2% (0.1-4.2) |
| Clipping nails | | | | 1.2% (0.1-4.1) | 1.2% (0.1-4.2) |
| Clinical Examination | 4.4% (0.5-15.2) | 9.7% (2.0-25.8) | 5.1% (0.6-17.3) | 4.6% (2.0-8.9) | 23.4% (17.3-30.5) |
| Clinical Procedure (unknown sedation) | | | | | 2.3% (0.6-5.9) |
| Clinical Procedure (with sedation) | 4.4% (0.5-15.2) | | 2.6% (0.1-13.5) | | 1.2% (0.1-4.2) |
| Feeding | | | 2.6% (0.1-13.5) | | |
| General petting | 2.2% (0.1-11.8) | 3.2% (0.1-16.7) | | | 2.3% (0.6-5.9) |
| Grooming | 2.2% (0.1-11.8) | | | 0.6% (0.0-3.2) | |
| Handling | 8.9% (2.5-21.2) | 3.2% (0.1-16.7) | 5.1% (0.6-17.3) | 4.0% (1.6-8.2) | 8.8% (5.0-14.1) |
| Injecting IV or IV cannula placement | 2.2% (0.1-11.8) | | 5.1% (0.6-17.3) | 3.5% (1.3-7.4) | 2.9% (1.0-6.7) |
| Injecting subcutaneously or intramuscularly | | | 2.6% (0.1-13.5) | | 2.9% (1.0-6.7) |
| Injecting (unknown route) | | | | | 1.2% (0.1-4.2) |
| Kennel cough vaccine administration | | | | | 0.6% (0.0-3.2) |
| Muzzling | | | 2.6% (0.1-13.5) | 2.9% (0.9-6.6) | 3.5% (1.3-7.5) |
| 'Out of nowhere' Attack | 2.2% (0.1-11.8) | 6.5% (0.8-21.4) | | 1.2% (0.1-4.1) | 4.7% (2.0-9.0) |
| Radiography | | | | 0.6% (0.0-3.2) | 1.2% (0.1-4.2) |
| Restraint | 11.1% (3.7-24.1) | 16.1% (5.5-33.7) | 25.6% (13.0-42.1) | 32.4% (25.5-39.9) | 4.1% (1.7-8.3) |
| Retrieving-Inserting from patient accommodation | 4.4% (0.5-15.2) | 3.2% (0.1-16.7) | 10.3% (2.9-24.2) | 4.6% (2.0-8.9) | 3.5% (1.3-7.5) |
| Retrieving or inserting animal from pet carrier | | | 2.6% (0.1-13.5) | 2.3% (0.6-5.8) | 2.9% (1.0-6.7) |
| Other | 2.2% (0.1-11.8) | 3.2% (0.1-16.7) | 2.6% (0.1-13.5) | | 1.2% (0.1-4.2) |
| Unknown | 2.2% (0.1-11.8) | | 2.6% (0.1-13.5) | 6.4% (3.2-11.1) | 5.3% (2.4-9.8) |
| **Non-animal injury** | **48.9% (33.7-64.2)** | **51.6% (33.1-69.9)** | **25.6% (13.0-42.1)** | **24.8% (18.6-32.0)** | **18.1% (12.7-24.7)** |
| Contact with electricity | | | | 0.6% (0.0-3.2) | |
| Contact with harmful substance | 4.4% (0.5-15.2) | | 2.6% (0.1-13.5) | 2.9% (0.9-6.6) | 1.2% (0.1-4.2) |
| Contact with machinery | | | | | 0.6% (0.0-3.2) |
| Cut by surgical sharp | | | | 0.6% (0.0-3.2) | 2.3% (0.6-5.9) |
| Cut by non-surgical sharp | 2.2% (0.1-11.8) | | | 2.3% (0.6-5.8) | 1.2% (0.1-4.2) |
| Exposure to fire | | | | 0.6% (0.0-3.2) | |
| Exposure to heat/steam | 2.2% (0.1-11.8) | 9.7% (2.0-25.8) | | | 1.2% (0.1-4.2) |
| Fall from same level | | | | 0.6% (0.0-3.2) | |
| Manual handling | 2.2% (0.1-11.8) | 3.2% (0.1-16.7) | 2.6% (0.1-13.5) | 4.6% (2.0-8.9) | 1.2% (0.1-4.2) |
| Needlestick | | | 10.3% (2.9-24.2) | 2.9% (0.9-6.6) | 7.0% (3.7-11.9) |
| Musculoskeletal | 2.2% (0.1-11.8) | | | | 0.6% (0.0-3.2) |

| | | | | | |
|---|---|---|---|---|---|
| Slips/Trips | 17.8% (8.9-32.1) | 25.8% (11.9-44.6) | | 6.9% (3.6-11.8) | 1.2% (0.1-4.2) |
| Struck by equipment/furniture | 15.6% (6.5-29.5) | 12.9% (3.6-29.8) | 7.7% (1.6-20.9) | 2.9% (0.9-6.6) | 1.8% (0.4-5.0) |
| Unknown | | | 2.6% (0.1-13.5) | | |

**S2 Mechanism and resultant injury of animal-related companion animal veterinary work-related injuries.**

| | | | Cat | | | | Dog | | | |
|---|---|---|---|---|---|---|---|---|---|---|
| | | | Veterinary Nurse | | Veterinary Surgeon | | Veterinary Nurse | | Veterinary Surgeon | |
| **Mechanism** | **Body part** | **Injury** | **Recent (n=91) (95% CI)** | **Severe (n=83) (95% CI)** | **Recent (n=68) (95% CI)** | **Severe (n=46) (95% CI)** | **Recent (n=41) (95% CI)** | **Severe (n=50) (95% CI)** | **Recent (n=49) (95% CI)** | **Severe (n=70) (95% CI)** |
| **Bite** | | | **62.6% (51.9-72.6)** | **69.9% (58.8-79.5)** | **63.2% (50.7-74.6)** | **82.6% (68.6-92.2)** | **73.2% (57.1-85.8)** | **68.0% (53.3-80.5)** | **75.6% (61.1-86.7)** | **82.9% (72.0-90.8)** |
| | *Abdomen* | | | | | | | | | |
| | | Puncture | 1.1% (0.0-6.0) | 1.2% (0.0-6.5) | | | | 2.0% (0.1-10.7) | | |
| | *Arm* | | | | | | | | | |
| | | Bruising | | | 1.5% (0.0-7.9) | | 4.9% (0.6-16.5) | | 6.1% (1.3-16.9) | 4.3% (0.9-12.0) |
| | | Laceration | 2.2% (0.3-7.7) | 1.2% (0.0-6.5) | 1.5% (0.0-7.9) | | 7.3% (1.5-19.9) | 4.0% (0.5-13.7) | 6.1% (1.3-16.9) | 5.7% (1.6-14.0) |
| | | Puncture | 9.9% (4.6-18.0) | 9.6% (4.3-18.1) | 8.8% (3.3-18.2) | 10.9% (3.6-23.6) | 14.6% (5.6-29.2) | 22.0% (11.5-36.0) | 8.2% (2.3-19.6) | 7.1% (2.4-15.9) |
| | *Foot* | | | | | | | | | |
| | | Laceration | | | | 2.2% (0.1-11.5) | | | | |
| | *Hand* | | | | | | | | | |
| | | Bruising | | 1.2% (0.0-6.5) | 2.9% (0.4-10.2) | 2.2% (0.1-11.5) | 9.8% (2.7-23.2) | 6.0% (1.3-16.6) | 18.4% (8.8-32.0) | 5.7% (1.6-14.0) |
| | | Degloving | | | | | | | | 5.7% (1.6-14.0) |
| | | Laceration | 2.2% (0.3-7.7) | 2.4% (0.3-8.4) | 2.9% (0.4-10.2) | | 7.3% (1.5-19.9) | 6.0% (1.3-16.6) | 14.3% (5.9-27.2) | 7.1% (2.4-15.9) |
| | | Puncture | 46.2% (35.6-56.9) | 53.0% (41.7-64.1) | 56.9% (42.2-70.7) | 60.9% (45.4-74.9) | 21.2% (10.6-37.6) | 18.0% (8.6-31.4) | 14.3% (5.9-27.2) | 32.9% (22.1-45.1) |
| | *Head* | | | | | | | | | |
| | | Bruising | | | | | 4.9% (0.6-16.5) | 2.0% (0.1-10.7) | 4.1% (0.5-14.0) | 1.4% (0.0-7.7) |
| | | Fracture | | | | | | | | 1.4% (0.0-7.7) |
| | | Laceration | | | | | | | | 5.7% (1.6-14.0) |
| | | Puncture | 1.1% (0.0-6.0) | 1.2% (0.0-6.5) | 1.5% (0.0-7.9) | 2.2% (0.1-11.5) | | 4.0% (0.5-13.7) | 4.1% (0.5-14.0) | 4.3% (0.9-12.0) |
| | *Leg* | | | | | | | | | |
| | | Bruising | | | | | | | | 1.4% (0.0-7.7) |
| | *Neck* | | | | | | | | | |
| | | Puncture | | | | | | 2.0% (0.1-10.7) | | |
| | *Multiple* | | | | | | | | | |
| | | Puncture | | 1.2% (0.0-6.5) | 1.5% (0.0-7.9) | 2.2% (0.1-11.5) | 2.4% (0.1-12.9) | 2.0% (0.1-10.7) | | |
| **Scratch** | | | **37.4% (27.4-48.1)** | **28.9% (19.5-39.9)** | **36.8% (25.4-49.3)** | **17.4% (7.8-31.4)** | **19.5% (8.8-34.9)** | **4.0% (0.5-13.7)** | **8.2% (2.3-19.6)** | **7.1% (2.4-15.9)** |
| | *Arm* | | | | | | | | | |
| | | Bruising | 1.1% (0.0-6.0) | 1.2% (0.0-6.5) | | 2.2% (0.1-11.5) | | | | |
| | | Laceration | 9.9% (4.6-18.0) | 8.4% (3.5-16.6) | 5.9% (1.6-14.4) | 6.5% (1.4-17.9) | 12.2% (4.1-26.2) | | 2.0% (0.1-10.9) | 2.9% (0.3-9.9) |
| | | Puncture | 2.2% (0.3-7.7) | | 5.9% (1.6-14.4) | | 4.9% (0.6-16.5) | 2.0% (0.1-10.7) | 2.0% (0.1-10.9) | |
| | *Back* | | | | | | | | | |
| | | Laceration | | | | | | | 2.0% (0.1-10.9) | 1.4% (0.0-7.7) |
| | *Hand* | | | | | | | | | |
| | | Laceration | 15.4% (8.7-24.5) | 9.6% (4.3-18.1) | 20.6% (11.7-32.1) | 6.5% (1.4-17.9) | 2.4% (0.1-12.9) | | | |
| | | Puncture | 4.4% (1.2-10.9) | 1.2% (0.0-6.5) | 2.9% (0.4-10.2) | 2.2% (0.1-11.5) | | | | |

| | | | | | | | | | | |
|---|---|---|---|---|---|---|---|---|---|---|
| | *Head* | Laceration | | 1.2% (0.0-6.5) | | | | | | |
| | *Leg* | Bruising | | | | | | 2.0% (0.1-10.7) | 2.0% (0.1-10.9) | 2.9% (0.3-9.9) |
| | | Laceration | | 1.2% (0.0-6.5) | | | | | | |
| | *Neck* | Puncture | 3.3% (0.7-9.3) | | | | | | | |
| | *Multiple* | Laceration | 1.1% (0.0-6.0) | 2.4% (0.3-8.4) | 1.5% (0.0-7.9) | | | | | |
| | | Puncture | | 1.2% (0.0-6.5) | | | | | | |
| **Ergonomic** | | | | | | | **4.9% (0.6-16.5)** | **14.0% (5.8-26.7)** | | **2.9% (0.3-9.9)** |
| | *Back* | Sprain | | | | | 2.4% (0.1-12.9) | 10.0% (3.3-21.8) | | 2.9% (0.3-9.9) |
| | *Hand* | Sprain | | | | | 2.4% (0.1-12.9) | 2.0% (0.1-10.7) | | |
| | *Head* | Concussion | | | | | | 2.0% (0.1-10.7) | | |
| **Headbutt** | | | | | | | **2.4% (0.1-12.9)** | **4.0% (0.5-13.7)** | **2.0% (0.1-10.9)** | |
| | *Head* | Bruising | | | | | 2.4% (0.1-12.9) | 4.0% (0.5-13.7) | | |
| | | Fracture | | | | | | | 2.0% (0.1-10.9) | |
| **Kick** | | | | | | | | | **2.0% (0.1-10.9)** | |
| | *Head* | Bruising | | | | | | | 2.0% (0.1-10.9) | |
| **Lead-related injury** | | | | | | | | **6.0% (1.3-16.6)** | **12.2% (4.6-24.8)** | **7.1% (2.4-15.9)** |
| | *Back* | Sprain | | | | | | 2.0% (0.1-10.7) | | |
| | *Foot* | Fracture | | | | | | | | 1.4% (0.0-7.7) |
| | *Hand* | Bruising | | | | | | | 4.1% (0.5-14.0) | 1.4% (0.0-7.7) |
| | | Fracture | | | | | | 2.0% (0.1-10.7) | 2.0% (0.1-10.9) | 1.4% (0.0-7.7) |
| | | Sprain | | | | | | | 2.0% (0.1-10.9) | |
| | *Shoulder* | Sprain | | | | | | 2.0% (0.1-10.7) | 4.1% (0.5-14.0) | 2.9% (0.3-9.9) |

**S3 – Consequences of severe work-related injuries in companion animal veterinary practices**

†These sections won't always add to 100% as multiple people could be present at an injury

| | Administrative staff (n=45) (95% CI) | Reception Staff (n=31) (95% CI) | Animal Care Assistants (n=39) (95% CI) | Veterinary Nurses (n=173) (95% CI) | Veterinary Surgeons (n=171) (95% CI) |
|---|---|---|---|---|---|
| **Was anyone else present at point of injury? †** | | | | | |
| No | 28.9% (16.4-44.3) | 29.0% (14.2-48.0) | 17.9% (7.5-33.5) | 18.5% (13.0-25.1) | 11.7% (7.3-17.5) |
| Admin | 2.2% (0.1-11.8) | | 2.6% (0.1-13.5) | 2.3% (0.6-5.8) | 1.8% (0.4-5.0) |
| Animal Care Assistant | | 3.2% (0.1-16.7) | | 0.6% (0.0-3.2) | |
| Owner or client | 13.3% (5.1-26.8) | 12.9% (3.6-29.8) | 5.1% (0.6-17.3) | 4.0% (1.6-8.2) | 46.8% (39.1-54.6) |
| Receptionist | 6.7% (1.4-18.3) | 12.9% (3.6-29.8) | 2.6% (0.1-13.5) | 1.7% (0.4-5.0) | 2.3% (0.6-5.9) |
| Student Vet Nurse | 4.4% (0.5-15.2) | | | 3.5% (1.3-7.4) | 1.8% (0.4-5.0) |
| Vet Nurse | 15.6% (6.5-29.5) | 25.8% (11.9-44.6) | 30.8% (17.0-47.6) | 24.3% (18.1-31.4) | 33.9% (26.9-41.5) |
| Vet Student | 2.2% (0.1-11.8) | | | 0.6% (0.0-3.2) | 0.6% (0.0-3.2) |
| Vet Surgeon | 24.4% (12.9-39.5) | 19.4% (7.5-37.5) | 48.7% (32.4-65.2) | 51.4% (43.7-59.1) | 8.2% (4.5-13.4) |
| Other | 4.4% (0.5-15.2) | | 2.6% (0.1-13.5) | 1.7% (0.4-5.0) | 0.6% (0.0-3.2) |
| **Medical Treatment** | | | | | |
| None | 8.9% (2.5-21.2) | 16.1% (5.5-33.7) | 7.7% (1.6-20.9) | 4.6% (2.0-8.9) | 12.9% (8.2-18.8) |
| Self-administered first aid | 46.7% (31.7-62.1) | 25.8% (11.9-44.6) | 38.5% (23.4-55.4) | 30.0% (23.3-37.5) | 33.3% (26.3-40.9) |
| First aid received | 4.4% (0.5-15.2) | 19.4% (7.5-37.5) | 7.7% (1.6-20.9) | 8.1% (4.5-13.2) | 6.4% (3.3-11.2) |
| Primary Care | 6.7% (1.4-18.3) | 12.9% (3.6-29.8) | 12.8% (4.3-27.4) | 20.2% (14.5-27.0) | 12.9% (8.2-18.8) |
| Minor Injury Unit | 11.1% (3.7-24.1) | 22.6% (9.6-41.1) | 25.6% (13.0-42.1) | 18.5% (13.0-25.1) | 19.9% (14.2-26.7) |
| Emergency Department | 15.6% (6.5-29.5) | 3.2% (0.1-16.7) | 5.1% (0.6-17.3) | 12.7% (8.1-18.6) | 11.1% (6.8-16.8) |
| Hospital Admission | 6.7% (1.4-18.3) | | 2.6% (0.1-13.5) | 4.0% (1.6-8.2) | 3.5% (1.3-7.5) |
| Hospital Outpatients | | | | 1.7% (0.4-5.0) | |
| Physiotherapist | | | | | |
| **Physical Recovery Time** | | | | | |
| < 7 days | 57.8% (42.2-72.3) | 54.8% (36.0-72.7) | 71.8% (55.1-85.0) | 35.8% (28.7-43.5) | 42.7% (35.2-50.5) |
| 7-14 days | 11.1% (3.7-24.1) | 25.8% (11.9-44.6) | 17.9% (7.5-33.5) | 42.8% (35.3-50.5) | 34.5% (27.4-42.1) |
| 15-28 days | 11.1% (3.7-24.1) | 3.2% (0.1-16.7) | 7.7% (1.6-20.9) | 9.2% (5.4-14.6) | 14.0% (9.2-20.2) |
| >28 days | 20.0% (9.6-34.6) | 16.1% (5.5-33.7) | 2.6% (0.1-13.5) | 12.1% (7.7-18.0) | 8.8% (5.0-14.1) |
| **Time off work** | | | | | |
| None | 75.6% (60.5-87.1) | 90.3% (74.3-98.0) | 79.5% (63.5-90.7) | 71.7% (64.3-78.3) | 79.5% (72.7-85.3) |
| <7 days | 11.1% (3.7-24.1) | | 17.9% (7.5-33.5) | 17.9% (12.5-24.5) | 15.8% (10.7-22.1) |
| 7-14 days | 4.4% (0.5-15.2) | 3.2% (0.1-16.7) | | 6.4% (3.2-11.1) | 1.8% (0.4-5.0) |
| 15-28 days | 2.2% (0.1-11.8) | | | 1.7% (0.4-5.0) | 2.9% (1.0-6.7) |
| >28 days | 6.7% (1.4-18.3) | 6.5% (0.8-21.4) | 2.6% (0.1-13.5) | 2.3% (0.6-5.8) | |
| **Was there a practice policy to minimise risk?** | | | | | |
| Yes | 31.1% (18.2-46.7) | 16.1% (5.5-33.7) | 30.8% (17.0-47.6) | 14.5% (9.6-10.6) | 7.0% (3.7-11.9) |
| No/Not aware | 68.9% (53.4-81.8) | 83.9% (66.3-94.6) | 69.2% (52.4-83.0) | 85.5% (79.4-90.4) | 93.0% (88.1-96.3) |
| **Did you inform practice management?** | | | | | |
| Yes | 86.7% (73.2-95.0) | 83.9% (66.3-94.6) | 94.9% (82.7-99.4) | 93.6% (88.9-96.8) | 82.5% (75.9-97.8) |
| **Did you file an accident/incident report?** | | | | | |
| Yes | 53.3% (37.9-68.3) | 71.0% (52.0-85.8) | 89.7% (75.8-97.1) | 82.5% (75.9-87.8) | 46.2% (38.6-54.0) |
| **Was any subsequent action taken by the practice?** | | | | | |
| Yes | 24.4% (12.9-39.5) | 9.7% (2.0-25.8) | 15.4% (5.9-30.5) | 16.8% (11.5-23.2) | 7.0% (3.7-11.9) |
| No | 60.0% (44.3-74.3) | 64.5% (45.4-80.8) | 64.1-47.2-78.8) | 59.0% (51.2-66.4) | 70.2% (62.7-76.9) |
| Don't know | 15.6% (6.5-29.5) | 25.8% (11.9-44.6) | 20.5% (9.3-36.5) | 24.2% (18.1-31.4) | 22.8% (16.8-29.8) |

**S4 Reasons for behavioural responses to a severe work-related injury in companion animal veterinary practices**

| | Administrators | Receptionists | Animal Care Assistants | Veterinary Nurses | Veterinary Surgeons |
|---|---|---|---|---|---|
| Did you experience any **emotional or mental** effects resultant of the injury? | n=45 | n=31 | n=39 | n=173 | n=171 |
| None | 84.4% | 87.1% | 89.7% | 75.7% | 76.6% |
| Increased anxiety or fear | 4.4% | 6.5% | 10.3% | 16.2% | 17.0% |
| More cautious around the animal | 6.7% | 3.2% | | 6.9% | 3.5% |
| Won't work with that species again | 2.2% | | | | 1.8% |
| Confidence knocked | | | | 1.2% | |
| Embarrassment | | 3.2% | | 0.6% | |
| Annoyance | | | | 0.6% | 1.8% |
| Undervalued by employer | 2.2% | | | 1.7% | 0.6% |
| Why did you **not take time off** from work? | n=34 | n=28 | n=31 | n=124 | n=136 |
| Genuinely minor injury not requiring time off work | 88.2% | 96.4% | 93.5% | 85.5% | 69.1% |
| Could still function with a reduced workload | 5.9% | | | 10.5% | 11.0% |
| 'Be tough'/'Just get on with it!' | 2.9% | | 3.2% | 3.2% | 4.4% |
| Don't want to lose sick leave-sick pay etc | | 3.6% | | 0.8% | |
| Not allowed to by employer | | | | 0.8% | 4.4% |
| Used annual leave | 2.9% | | 3.2% | | 2.2% |
| Didn't want to let team down | 2.9% | | | 0.8% | 12.5% |
| Why did you choose **not to report** the incident? | n=19 | n=9 | n=4 | n=26 | n=76 |
| Only minor injury, which wasn't serious enough | 21.1% | 66.7% | 50.0% | 38.5% | 31.6% |
| Unaware it needed reporting | 5.3% | | | 7.7% | 10.5% |
| Didn't want to make a fuss | | | | 3.8% | 1.3% |
| Too much effort as was too busy | | 22.2% | | 15.4% | 10.5% |
| It would cause tension amongst the team | | | | 3.8% | |
| It's just one of those things/'par for the course' | 5.3% | | 25.0% | 19.2% | 13.2% |
| Not employed at CVS at the time | 31.6% | 22.2% | 25.0% | 3.8% | 2.6% |
| I forgot to do it | 10.5% | | | 19.2% | 9.2% |
| No reporting system in place | 26.3% | | | 3.8% | 32.9% |
| Have you **changed your behaviour** around the animal species involved in the injury? | n=41 | n=29 | n=38 | n=167 | n=168 |
| No | 46.3% | 51.7% | 39.5% | 35.3% | 29.8% |
| More cautious and less confident | 7.3% | 27.6% | 13.2% | 10.2% | 19.6% |
| Avoidance of procedure or animal | 7.3% | | 5.3% | 3.6% | 7.7% |
| Do procedure differently | 26.8% | 13.8% | 7.9% | 18.0% | 23.8% |
| Improve handling/restraint | 4.9% | 3.4% | 23.7% | 16.2% | 12.5% |
| Would now use PPE-Muzzle | 12.2% | 3.4% | 7.9% | 12.0% | 16.7% |
| Denial that did anything wrong | | | 2.6% | 3.0% | 4.8% |
| Improved awareness of surroundings | 2.4% | 10.3% | | 6.0% | 3.6% |

### Supportive Quotes:

Increased anxiety of fear

*"I feared taking time off would cause me to become more fearful and that the best way to over it was to get right back into it. I had to remain on light duties as was only able to use one hand for a week or so"* – VN dog bite to hand, attended hospital emergency department

Reduced workload

*"I was able to do admin work but not clinical work. I was fit for some clinical work after 2-3 days" – VS, cat bite to hand, received first aid*

‘Be tough’/’Just get on with it!’

*“Didn't feel like I needed to. It would take a lot for me to have time off”. – VS, cat bite to hand, attendance at primary care physician*

Too much effort as was too busy to report an injury

*“Too much to do, too little time in which to do it. Work load is often overwhelming so optional things like reporting minor injuries get neglected. “*– VS, cat scratch to hand, received first aid

It’s just one of those things/’par for the course’

*“Common occurrence/hazard in a veterinary practice and not concerned.”* – VS, cat scratch to hand, received first aid

‘More cautious, and less confident’

*“Bit more wary of even apparently fully sedated dogs!”* – VS, dog bite to hand, attendance at hospital emergency department.

Do procedure differently

*“Will make sure to give heads up to anyone holding the animal as giving injection and ask for others to do the same if I am holding”* – VS, dog bite arm, received first aid

Improve restraint techniques

*“I usually hold fractious cats myself now and struggle to trust others to hold for me.”* – VN, cat bite to hand, hospital admission

*“Much quicker to postpone manual restraint and instead request the animal returns with some level of chemical restraint present”* – VS, hand crushed by dog, no medical treatment

Would now muzzle

*"All risk animals are muzzled and muzzles are not removed whilst there is a risk to persons involved."*-

VN, dog bites to hands and arms, attendance at emergency department
